# Decomposing the genetic background of chronic back pain

**DOI:** 10.1101/2024.02.14.24302763

**Authors:** Elizaveta E. Elgaeva, Irina V. Zorkoltseva, Arina V. Nostaeva, Dmitrii A. Verzun, Evgeny S. Tiys, Anna N. Timoshchuk, Anatoliy V. Kirichenko, Gulnara R. Svishcheva, Maxim B. Freidin, Frances M. K. Williams, Pradeep Suri, Yurii S. Aulchenko, Tatiana I. Axenovich, Yakov A. Tsepilov

## Abstract

Chronic back pain (CBP) is a disabling condition with a lifetime prevalence of 40% and a substantial socioeconomic burden. Because of the high heterogeneity of CBP, subphenotyping may be necessary to improve prediction and support personalized treatment for those with CBP. The lack of distinct cellular and molecular markers for CBP complicates the task of subphenotyping.

To investigate CBP subphenotypes, we decomposed the genetic background of CBP into a shared genetic background common to other chronic pain conditions (back, neck, hip, knee, stomach, and head pain) and unshared genetic background related only to CBP. We showed that the shared and unshared genetic backgrounds of CBP differ in their biological functions: the first one is likely to control processes mainly in nervous, immune and musculoskeletal systems underlying chronic pain development regardless its site, while the second may contribute more to local processes in spine leading to chronic pain precisely in the back. We identified 18 genes with shared impact across different chronic pain conditions and two genes that were specific for CBP. These findings may contribute to future development of targets and new biomarkers for chronic pain management.

Next, among people with CBP, we demonstrated that polygenic risk scores accounting for the shared and unshared genetic backgrounds of CBP may underpin different subphenotypes of CBP cases. These subphenotypes are characterized by varying genetic predisposition to a wide array of medical conditions and interventions such as diabetes mellitus, myocardial infarction, diagnostic endoscopic procedures, and surgery involving muscles, bones, and joints. The proposed genetic decomposition framework holds promise for investigating the genetic underpinnings of other heterogeneous diseases.

**Author Summary:** Chronic back pain (CBP) is a prevalent disabling health problem with heterogeneous clinical presentation and natural history. This may contribute to generic pain treatment approaches not sufficiently effective when prescribed for patients with certain characteristics. Development of more personalized treatment is needed, and may benefit from a deep understanding of CBP biology, such as genomics. It is known that chronic pain is under the control of both environmental and genetic background. Here we applied bioinformatic methods to study the genetic background of CBP decomposed into two parts: a shared one common to six distinct chronic pain types, and unshared, which is specific to CBP. This approach allowed us to identify more genes potentially involved in CBP development. Among them 18 belong to the shared genetic background contributing to development of chronic pain in general, and two are specific for CBP. We demonstrate that these two parts of the genetic background of CBP are associated with distinct biological pathways and underlie predisposition to different medical states and procedures, involving diabetes, myocardial infarction, and musculoskeletal surgery. Decomposition of CBP genetic background into shared and unshared may provide a better understanding of mechanisms of CBP and facilitate development of personalized pain treatment.

## Introduction

Back pain is a prevalent clinical syndrome which affects about 40% of the population [1]. It has tremendous social and economic consequences: according to the Global Burden of Disease Study 2016, back pain has been a major cause of disability worldwide for 30 years [2]. Back pain is not only highly prevalent, but it is also difficult to treat [3]. One potential cause of this problem is the high heterogeneity of the condition. Patients with a specific back pain “subphenotype” – a set of common features, distinguishing them from other patients with back pain [4] – may respond to treatment in a different way than a patient of another back pain subphenotype, decreasing the effectiveness of treatment approaches when not tailored to subphenotype. Generic treatment approaches may contribute to back pain patient care costs, which reach 1/5 of total health care costs in a separate country [5]. Subphenotyping may help to more accurately select treatment for patients with back pain and thus decrease these costs.

In 10% of cases acute back pain ceases to be just a symptom and becomes a chronic condition [6]. While the initial cause of acute pain may resolve, an alternative pathophysiological process takes over, leading to anatomical changes and affecting human behavior and mental state [7–9]. Chronic back pain (CBP) has been shown to be a complex trait with heritability estimated between 30 and 68% [10–12]. Genome-wide association studies (GWAS) of chronic pain including but not limited to CBP have revealed about three dozen associated loci [12–17], but only a few of them have been replicated in independent samples.

Studies have demonstrated the presence of a shared genetic background of chronic pain across different pain sites [16,18,19]. This shared genetic background is thought to condition the generation, transduction and processing of pain stimuli in general [16]. At the same time, it is reasonable to expect the existence of unshared genetic background specific to chronic back pain (CBP) and related to local pathological processes in the back and spine (e. g. through *SOX5* gene [15]), but this has been little studied. Genetic variants related to unshared genetic background of CBP may suggest promising drug targets and biomarkers for diagnosis and treatment of CBP precisely, while the shared genes could be of interest to understand general pain biology and hence contribute in future development of drugs to treat pain irrespective of its site. Therefore, the decomposition of genetic background into shared and unshared may be helpful for subphenotyping and personalizing treatment.

Many prior attempts to identify subphenotypes of back pain have been made. Existing approaches to do this classify patients based on observable clinical characteristics such as pain-related, social, physiological and anatomic features [20] and some of these studies show the utility of this approach for revealing clusters of patients with specific pain trajectories and differentially reacting to treatment [21,22]. A novel and alternative approach is to subphenotype CBP using genotypic information, which is increasingly available in clinical care and commercial use [23]. Polygenic risk scores (PRS) use GWAS data to estimate the personal genetic liability to disease based on an individual’s genotype. In medicine, PRSs have been used to predict disease, however, this information can also be utilized for subphenotyping. While existing chronic back pain PRS models show modest prediction ability [24], division of CBP genetic background into shared and unshared, followed by PRS calculation for each of them, may result in higher statistical power and provide useful information for patients’ subphenotyping. To estimate the potential of genetic background decomposition for subphenotyping, a comprehensive examination of the features of shared and unshared genetic background is required.

Previously, we have studied the shared genetic background of chronic musculoskeletal pain at different sites (back, neck, hip, knee) by applying principal component analysis of these traits [16]. We interpreted the first genetically independent phenotype (GIP1, or simply the first principal component) as an approximation of shared genetic background across the considered pain types. In our recent study we developed the novel SHAHER framework, [25] allowing more accurate decomposition of genetic background of correlated traits in order to evaluate not only their shared, but unshared genetic factors as well. Here we aimed to employ SHAHER to investigate the complex genetic architecture of CBP by analyzing its shared genetic background across several chronic musculoskeletal and non-musculoskeletal pain conditions (back, neck, hip, knee, stomach and head pain) and its unshared genetic background, particular to chronic pain in the back. We conducted an extensive bioinformatic annotation of shared and unshared genetic background of CBP to reveal functional differences and identify genes associated with each aspect. To find the associated genes, we additionally performed a gene-based association analysis. Finally, we performed a set of PRS analyses among people with CBP to check whether the division between shared and unshared genetic background might be beneficial for CBP subphenotyping.

## Results

### Overview of the study design

This study was carried out using the results of genome-wide association studies (GWAS) for six chronic pain sites (pain in the back, neck, hip, knee, stomach, and headache). All data (Ntotal = 456,000) were provided by UK Biobank under projects #18219 and #59345. The sample was split into discovery (265,000 individuals of European descent) and replication (a total of 191,000 individuals in three subsamples of African, South Asian and European descent) samples. Details of the phenotypes definition are available in Supplementary Methods. Sample characteristics (size, sex and age structure, pain type prevalence and BMI distribution) are provided in Supplementary Table 1.

The study design included four stages. In the first stage, we decomposed the genetic background of CBP into shared and unshared ones, and calculated the GWAS summary statistics for two new traits: SGIT (shared genetic impact trait) which is controlled predominantly by shared genetic background common for all traits analyzed, and UGIT (unshared genetic impact trait) which is controlled by genetic background specific for CBP. These calculations were performed for each subsample using the SHAHER framework (Figure 1, Materials and Methods). Then we performed two meta-analyses for SGIT and UGIT using inverse-variance-weighted method: one combining the summary statistics from the two European samples (European meta-analysis) and another one combining summary statistics from the three replication samples (Replication meta-analysis). Thus, four sets of GWAS summary statistics both for SGIT and UGIT were obtained in the first stage: the “Discovery sample”, the “European replication sample”, the “European meta-analysis” and the “Replication meta-analysis” (Figure 1).

**Figure 1.**
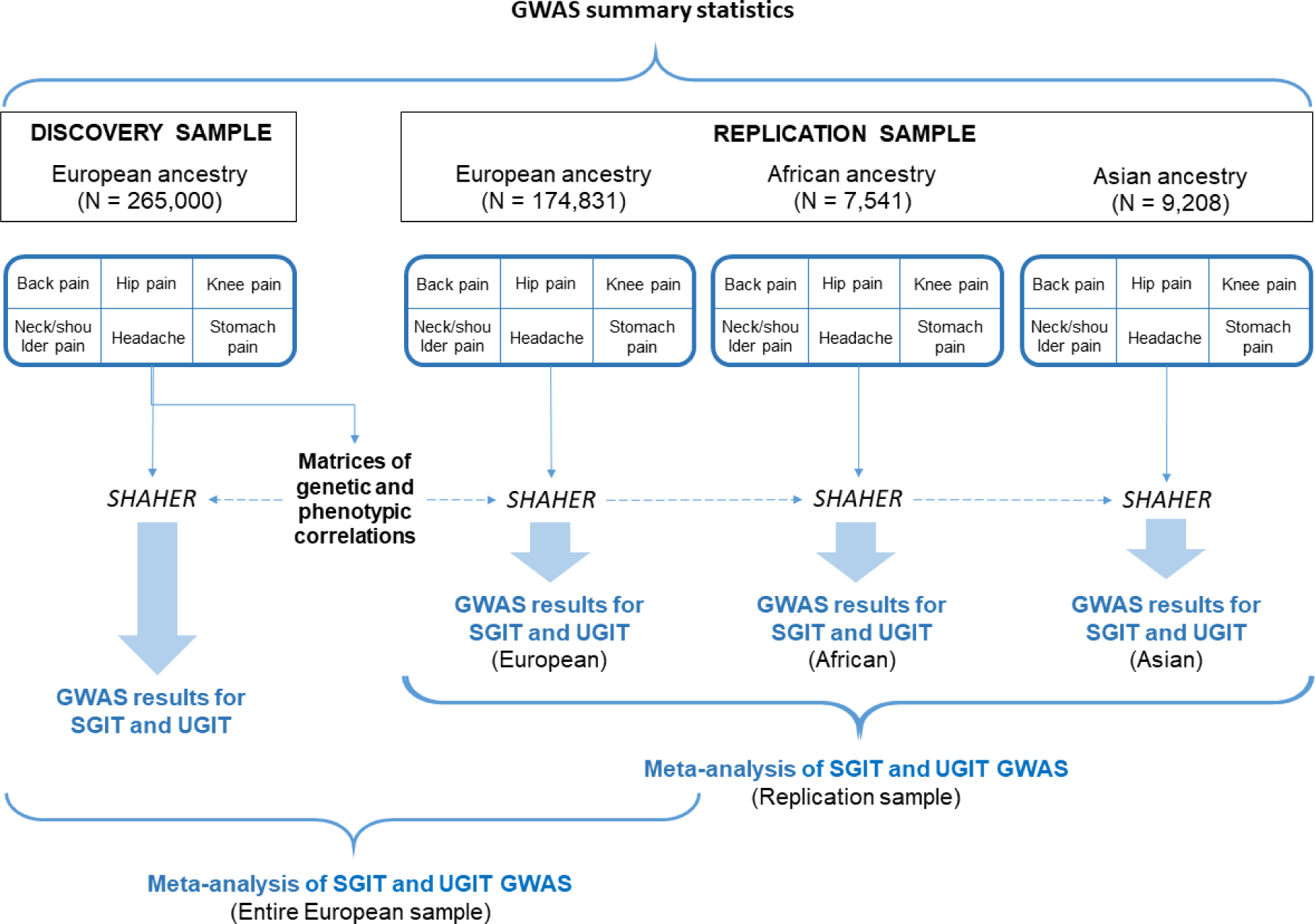
The first step of the study. The black frames label the analyzed sample and the blue frames indicate the input data; the black text in bold shows intermediate results; the text written in blue bold highlights the output data, and the black text written in italic represents the type of analysis.

In the second stage, we identified loci and genes associated with SGIT and UGIT. We did this using two approaches. The first approach (Figure 2) was identification of associated loci in the discovery sample, followed by selection of independent association signals within these loci utilizing conditional analysis. For those loci that were previously observed and replicated in our recent work utilizing the same data [15,16], we did not perform additional analyses. For other association signals, we conducted replication using the Replication meta-analysis. For loci replicated in the Replication meta-analysis we carried out gene prioritization procedure using the European meta-analysis. Gene prioritization included prediction of SNP effects in replicated loci (VEP and FATHMM), analysis of colocalization with gene expression effects (SMR-HEIDI), and gene prioritization using DEPICT and FUMA. Details are available in Supplementary Methods. The second approach utilizing which we identified genetic factors associated with shared and unshared backgrounds of CBP (Figure 2) was a gene-based association analysis (SKAT-O [26], PCA [27], ACAT-V [28], and ACAT-O [28] methods) using the GWAS summary statistics obtained from the discovery sample, followed by a conditional analysis and replication utilizing the gene-based association analysis results from the European replication sample and European meta-analysis (Figure 2, Materials and Methods).

**Figure 2.**
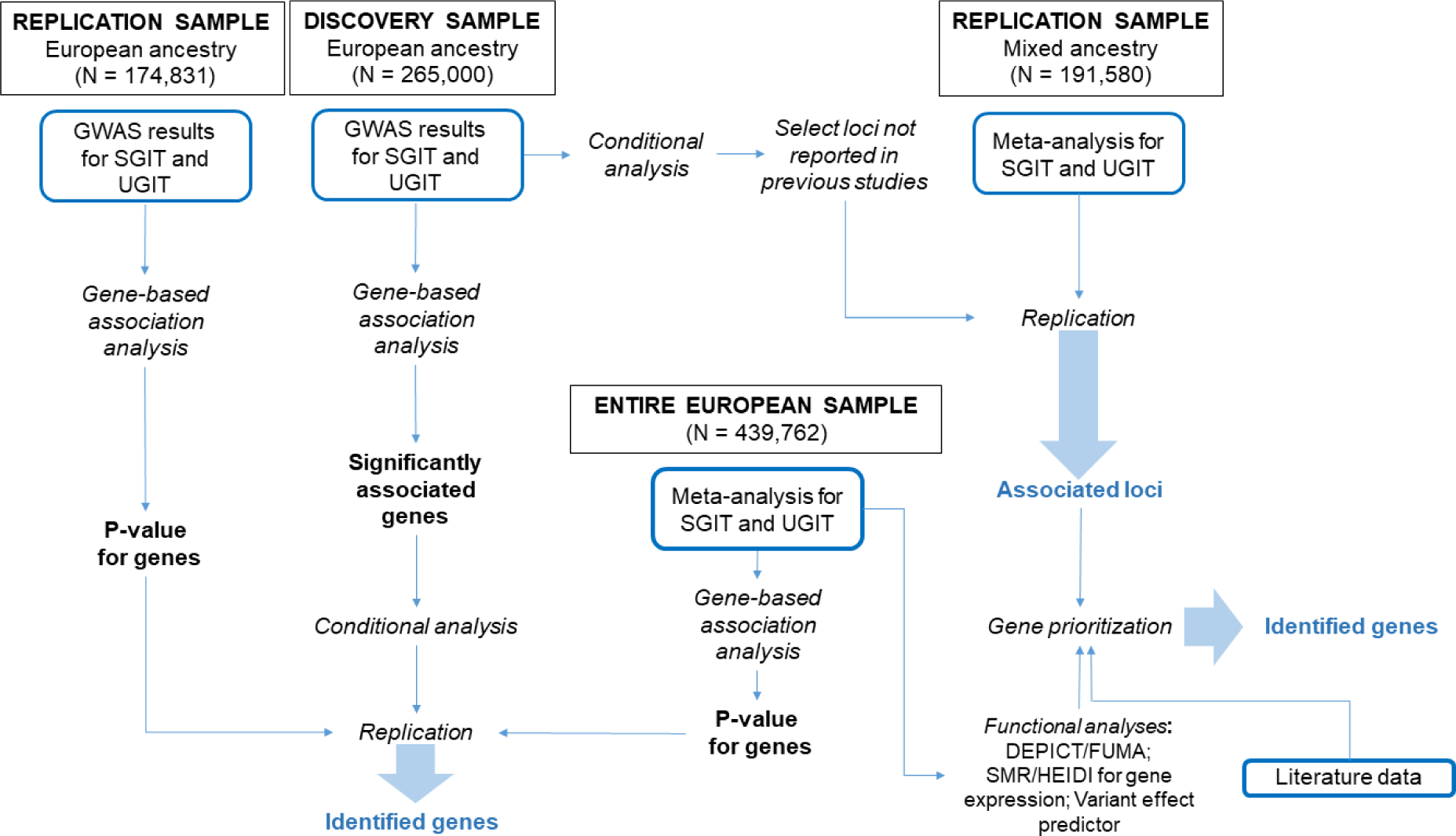
The second step of the study. The black frames label the analyzed sample and the blue frames indicate the input data; the black text in bold shows intermediate results; the text written in blue bold highlights the output data, and the black text written in italic represents the type of analysis.

At the third stage (Figure 3), we investigated the genetic architecture of CBP using functional bioinformatic analyses. We conducted a gene set and cell type/tissue enrichment analyses of the identified genes and SNPs in the associated loci in order to characterize the shared and unshared genetic background of CBP. Using GWAS summary statistics obtained for SGIT and UGIT, we estimated the genetic correlations of these traits with a large number of complex human traits from the GWAS-MAP database [29,30]. Then we calculated polygenic risk scores (PRS) of SGIT and UGIT using individual genotype data and estimated their role in disease/medical intervention prediction.

**Figure 3.**
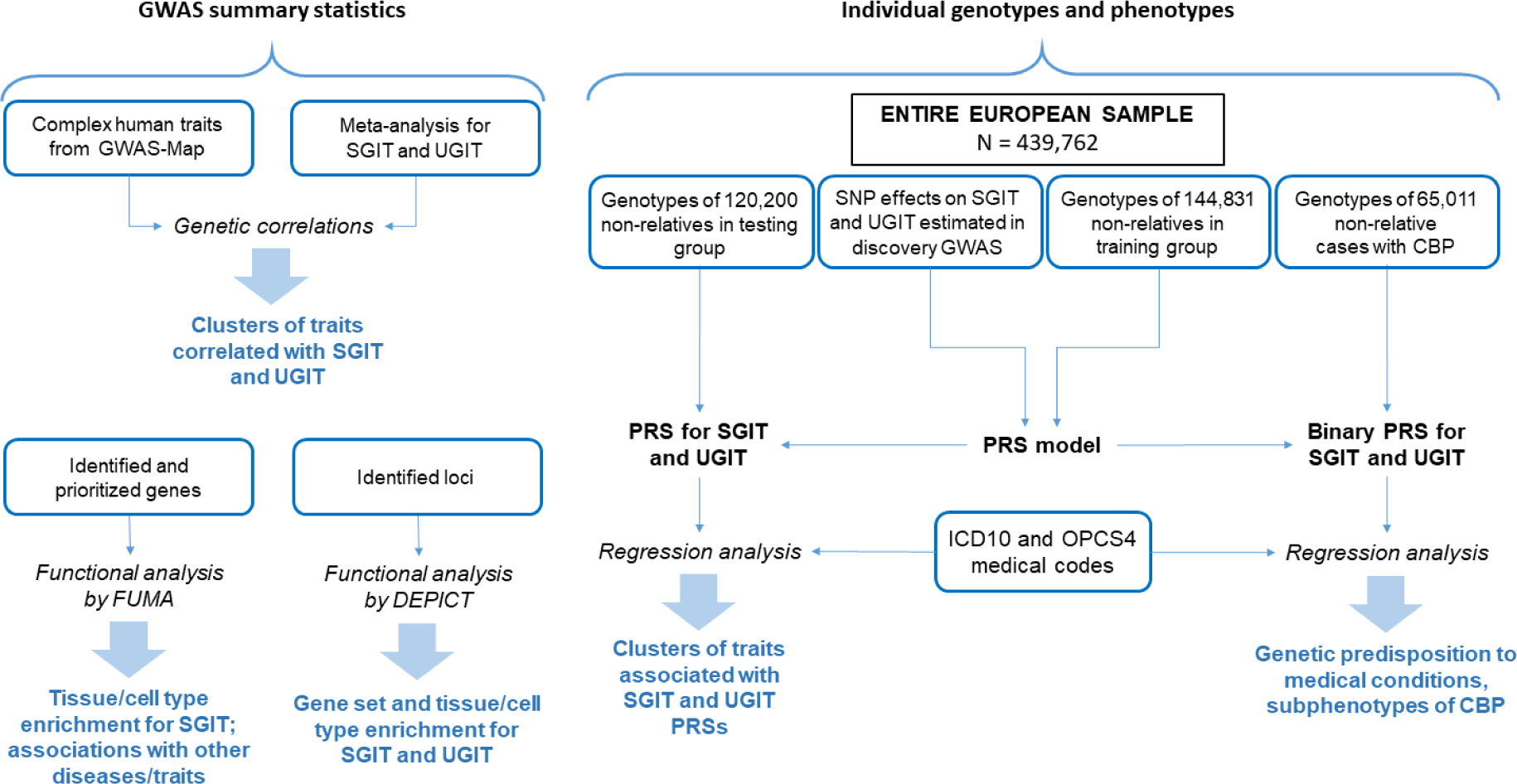
The third and the fourth steps of the study. The black frames label the analyzed sample and the blue frames indicate the input data; the black text in bold shows intermediate results; the text written in blue bold highlights the output data, and the black text written in italic represents the type of analysis.

Finally, we investigated the potential of SGIT and UGIT PRSs for CBP subphenotyping (Figure 3, Materials and Methods) using participants with CBP from the entire European sample (not divided into discovery and replication). We built binary-coded PRSs for SGIT and UGIT by splitting the PRS distribution into two parts according to the lowest or highest decile (see Materials and Methods). Then we tested if the binary-coded PRSs can be used for CBP subphenotyping.

### Decomposition of genetic background of CBP

We estimated the heritability, the genetic correlation and phenotypic correlation for six chronic pain traits from the discovery sample (see Supplementary Methods and Supplementary Tables ST2a, b). Using these estimates and applying the SHAHER framework, we calculated the coefficients of optimal linear combinations for SGIT and UGIT building (Supplementary Table ST2a) and obtained the summary statistics for SGIT and UGIT. Heritability estimates and correlation coefficients of the original pain traits, SGIT and UGIT are depicted in Figure 4. Notably, the SNP-based heritability of SGIT is almost two times higher than those of the original traits (0.07 versus 0.01 – 0.04). The phenotypic and genetic correlations between SGIT and the original traits are higher in comparison to those between the original traits. UGIT shows positive correlation only with CBP.

**Figure 4.**
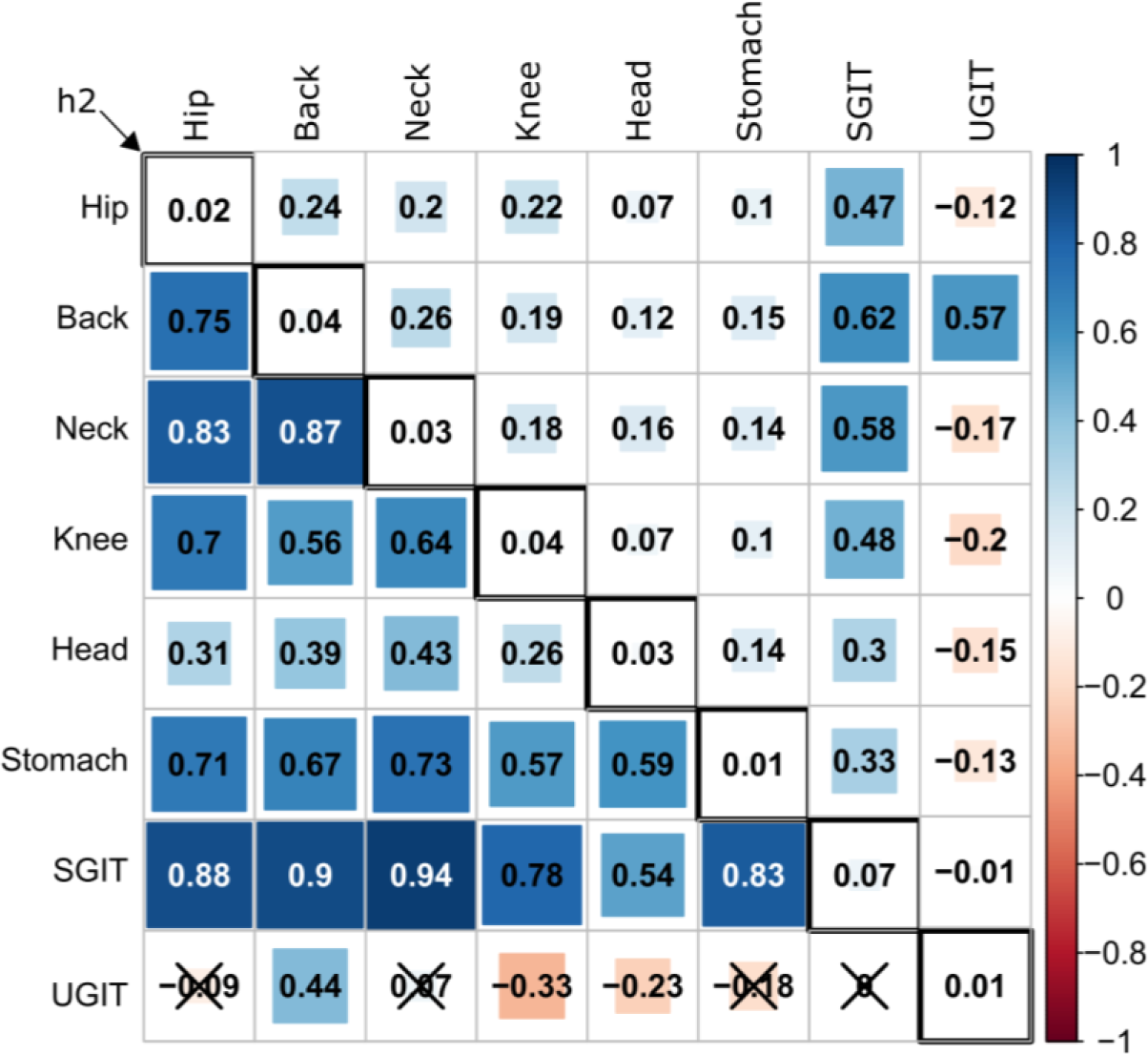
Heatmap visualization of the heritability, phenotypic correlations and genetic correlations of the original pain traits, SGIT and UGIT. Inscriptions name anatomic sites of chronic pain, UGIT refers to unshared genetic background of CBP. The upper triangle of the matrix represents pairwise phenotypic correlations. The lower triangular of the matrix contains pairwise genetic correlations. Statistically insignificant (p-value > 0.05/28) genetic correlation coefficients are crossed out. Diagonal elements of the matrix correspond to SNP-based heritability (h^2^) estimates of the particular traits.

### Gene identification

#### Identification of loci associated with SGIT and UGIT

The results of SGIT and UGIT GWASs obtained from the discovery sample are visualized at Manhattan and QQ plots (Supplementary.Figures 1 – 4). We identified five loci statistically significantly (p-value < 8.3e-09) associated with SGIT (see Table 1). Conditional analysis showed that each of these loci contains only one independent signal. Four loci (tagged with the following lead SNPs: rs9436127, rs7652179, rs12705966 and rs73581580) were previously observed and replicated in our recent study [16] in the first genetically independent phenotype (GIP1). The locus with the corresponding lead SNP rs11079993 was the new one. Similarly, we identified one locus with rs1271351 as the leading SNP significantly associated with UGIT. Conditional analysis showed a single independent association signal in this region. Since this locus has been already reported in [15], we interpreted it as known. Detailed information on significant loci and their replication is available in Supplementary Table ST3.

**Table 1.** Loci associated with SGIT and UGIT.

| Trait | Lead SNP | Chr:position | RefA/<br>EffA | Nearest<br>gene | Discovery sample |  |  |  | Replication meta-analysis |  |  |
| --- | --- | --- | --- | --- | --- | --- | --- | --- | --- | --- | --- |
| | | | | | $\beta$ | se | p-value | EAF | $\beta$ | se | p-value |
| SGIT | rs9436127 | 1:150490565 | G/A | *ECM1* | -0.018 | 0.003 | 1.72e-10 | 0.40 | -0.012 | 0.003 | 5.98e-04 |
|  | rs7652179 | 3:49808618 | T/A | *AMIGO3* | 0.023 | 0.004 | 2.30e-10 | 0.18 | 0.011 | 0.004 | 1.00e-02 |
|  | rs12705966 | 7:114248851 | G/A | *FOXP2* | 0.017 | 0.003 | 6.55e-09 | 0.67 | 0.011 | 0.004 | 2.16e-03 |
|  | rs73581580 | 9:140251458 | G/A | *EXD3* | 0.026 | 0.004 | 1.29e-09 | 0.12 | 0.029 | 0.005 | 5.36e-09 |
|  | **rs11079993** | **17:50301552** | **T/G** | ***snoZ178*** | **-0.017** | **0.003** | **4.98e-09** | **0.62** | **-0.013** | **0.003** | **7.94e-05** |
| UGIT | rs1271351 | 10:73798873 | T/C | *CHST3* | -0.018 | 0.003 | 1.13e-10 | 0.43 | -0.014 | 0.003 | 3.40e-05 |

The new replicated locus is shown in bold. Associations shown in grey were observed in previous studies [15,16]. Lead SNP – the SNP with the lowest p-value in the locus; Chr:position – genomic coordinates in a format “chromosome number : base pairs” (according to the GRCh37 genomic build); RefA – reference (not effective) allele; EffA – effective allele; **β** – effect size of SNP counted for effective allele; se – standard error of **β**; p-value – p-value of association between SNP and a trait after correction for genomic control; EAF – effective allele frequency.

#### Prioritization of genes in the new associated locus

We conducted gene prioritization for the rs11079993 locus associated with SGIT utilizing several approaches: literature-based prioritization of the genes located in this locus; prediction of pathogenicity of SNP effects in the locus using VEP [31], FATHMM-XF [32] and FATHMM-INDEL [33]; DEPICT [34] and FUMA [35] gene prioritization and estimation of pleiotropic effects on gene expression using SMR-HEIDI [36]. Gene prioritization suggested two genes in the rs11079993 locus (see Supplementary Results and Supplementary tables ST4-6 for more details): *CA10* (prioritized by literature-based annotation and FUMA) and *LINC01982* (prioritized by literature-based annotation). Although the latter is well studied it provided less evidence for prioritization than *CA10*, so we consider it less likely to be causal. The *CA10* gene may have an effect on chronic pain through processes in the central nervous system, cancer or disease of bones. Despite the fact that *snoZ178* is the nearest gene to rs11079993, it did not provide strong arguments for being causal.

#### Gene-based association analysis

We performed the gene-based association analysis using different SNP annotation sets within genes. For SGIT we detected 9, 18, and 43 genome-wide significant gene-based signals for nonsynonymous, protein coding and protein non-coding SNP sets, respectively (Supplementary Table ST7). After conditional analysis, 10 out of 43 for non-coding SNP set and one for each coding and nonsynonymous SNP sets (both in *SLC39A8*) retained significance. We replicated five of 12 genes using the European replication sample and European meta-analysis (Table 2). For UGIT we detected two significant signals for the non-coding SNP set, and one of them (*CHST3*) passed conditional analysis and replication.

**Table 2.** Results of gene-based association analysis of SGIT and UGIT.

| Trait | Gene | Chr | Position | SNP set | Gene-based p-value |  |  |  |
| --- | --- | --- | --- | --- | --- | --- | --- | --- |
|  |  |  |  |  | Discovery sample | Conditional analysis | European replication sample | European meta-analysis |
| UGIT | *CHST3* | 10 | 71964395 | ncod | 7.81e-08 | 7.81e-08* | **1.42e-06** | **7.93e-13** |
| SGIT | *SLC39A8* | 4 | 102251080 | nsyn | 5.68e-10 | 5.68e-10* | **4.73e-09** | **4.38e-17** |
|  | *SLC39A8* | 4 | 102251080 | cod | 1.17e-07 | 1.17e-07* | **1.62e-06** | **2.54e-13** |
|  | *IP6K1* | 3 | 49724294 | ncod | 1.32e-10 | 1.32e-10* | 0.0083 | **2.66e-12** |
|  | *GRM3* | 7 | 86643909 | ncod | 1.66e-07 | 1.66e-07* | 0.0935 | **6.55e-08** |
|  | *FOXP2* | 7 | 114086327 | ncod | 2.00e-07 | 2.00e-07* | **7.90e-04** | **8.22e-11** |
|  | *MAML3* | 4 | 139716753 | ncod | 3.17e-07 | 3.17e-07* | **6.40e-05** | **1.30e-12** |
|  | *PTBP1* | 19 | 797075 | ncod | 3.64e-07 | 3.64e-07* | 0.8760 | **2.33e-07** |
|  | *TCF20* | 22 | 42160013 | ncod | 1.93e-06 | 1.93e-06* | **3.77e-04** | **8.44e-10** |
|  | *SOX6* | 11 | 15966449 | ncod | 2.01e-06 | 2.01e-06* | 0.0921 | **4.17e-07** |
|  | *FZD10* | 12 | 130162459 | ncod | 2.03e-06 | 2.22e-06 | 0.2626 | 4.36e-06 |
|  | *ERICH2* | 2 | 170766878 | ncod | 2.25e-06 | 2.25e-06* | 0.1304 | 3.19e-05 |
|  | *GABRB2* | 5 | 161288429 | ncod | 2.08e-06 | 2.40e-06 | **5.20e-05** | **3.69e-11** |

\* These genes harbored SNPs with the lowest p-values within 5 Mb from their borders, thus no conditional SNPs were selected, and the gene-based p-values remained unchanged.
The significance threshold for replication sample was set at $p\text{-value} < 4.2e-03 = 0.05/12$ , where 12 is the number of genes retained significance after the conditional analysis.

#### Summary of gene identification

Using prioritization of genes in the associated loci and gene-based association analysis, we identified 18 genes associated with SGIT, with 13 of them being previously reported in our recent works [15,16] (*SLC39A8, FOXP2, EСM1, AMIGO3, BSN, RBM6, FAM212A, RNF123, UBA7, MIR7114, NSMF, NOXA1, GRIN1*), and 5 of them being new (*CA10, MAML3, TCF20, GABRB2, LINC01982*). One of the identified genes, namely *FOXP2*, was found using both gene prioritization and gene-based analysis. The *SLC39A8* gene was identified using gene-based analysis and has been prioritized in our previous study using the genetically independent phenotype (GIP) approach [16].

For UGIT, we identified the *CHST3* gene utilizing gene-based analysis, and prioritized two genes in the UGIT-associated locus: *CHST3* and *SPOCK2,* failing to give preference to one of them [15].

### Analysis of CBP genetic architecture

#### Gene set and tissue/cell type enrichment

We performed two enrichment analyses based on DEPICT for SNPs associated with SGIT and UGIT in European meta-analysis, and FUMA analysis for the genes identified for SGIT. In DEPICT analysis at p-value < 5e-06 we found SGIT to be enriched in genes involved in the BTBD2 PPI subnetwork and decreased cochlear coiling (Supplementary Table ST8a). We also detected enrichment for genes expressed in the central nervous and neurosecretory systems, retina and neural stem cells (Supplementary Table ST8b). Similar analyses of SGIT-associated variants at p-value < 2.5e-08 threshold provided no significant findings. For UGIT we were able to conduct DEPICT analysis only for SNPs with p-value < 5e-06, but no statistically significant results were obtained. However, we observed a tendency to enrichment for genes expressed in the digestive, nervous, musculoskeletal, cardiovascular, hemic and immune systems.

We did not obtain statistically significant results in tissue specificity and gene set enrichment analyses using FUMA, however, we have shown that SGIT-associated genes have different expression patterns. For instance, there is a group of genes (*NSMF*, *PTBP1*, *RBM6*, *IP6K1*, *UBA7*, *ECM1*, *NOXA1*, *RNF123*) expressed almost in every organ, and a group of genes expressed predominantly in the brain (*GRIN1*, *GABRB2*, *GRM3*, *BSN*, and *CA10*) (Supplementary Figure 5). These findings are in line with those obtained for SGIT loci utilizing DEPICT. FUMA extends DEPICT findings for SGIT, by accounting for genes identified in gene-based analysis as well and providing more detailed information on expression of particular genes.

#### Genetic correlation between SGIT, UGIT and complex traits

We estimated the genetic correlation between each of SGIT and UGIT, and 730 complex human traits from the GWAS-Map database. SGIT was statistically significantly genetically correlated with almost all of the 322 preselected complex traits, and UGIT provided significant correlations with only 14 of them (see Supplementary Table ST9). In total we grouped all traits in 11 clusters (see Supplementary Table ST9, Figure 5). There were some distinctions between SGIT and UGIT correlation patterns. First, UGIT was significantly correlated with sitting height which represents the length of the spine, and the second, third and fourth genetically independent phenotypes (GIP2- 4) from our recent study [16], while SGIT was not (Supplementary Table ST9). Second, SGIT had a positive direction of genetic correlation with self-reported chronic knee pain and headache, when UGIT was negatively correlated with them. Regardless of statistical significance, SGIT and UGIT demonstrated opposite patterns of genetic correlation (see Figure 5). While SGIT was mostly positively correlated with traits from such clusters as: respiratory illness and smoking, injuries, socio-economic and family status, psychometric traits, osteoarthritis and other musculoskeletal disorders, UGIT tended to be inversely correlated with these clusters of traits.

**Figure 5.**
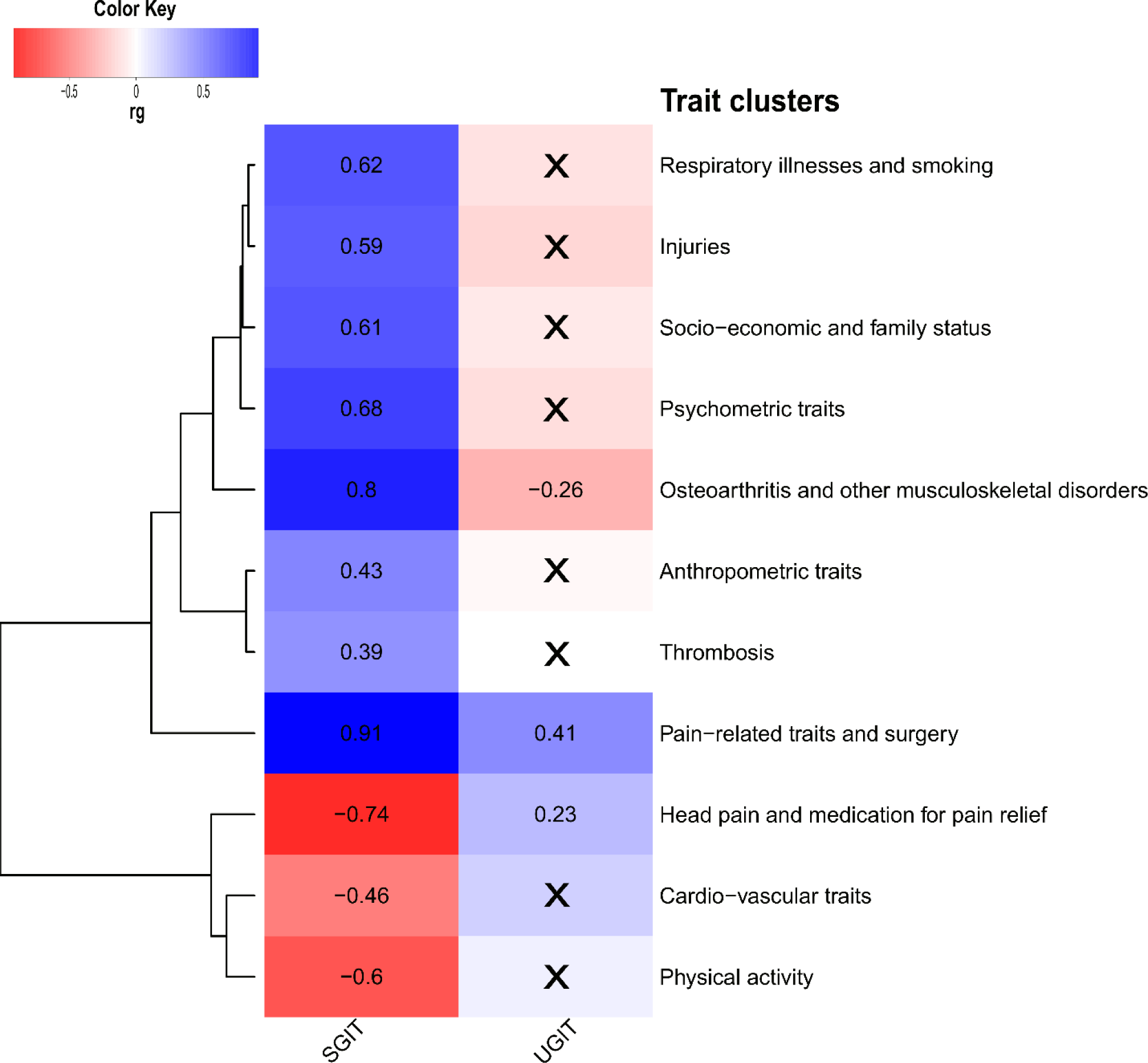
Genetic correlation of SGIT and UGIT with complex human traits. Only 322 traits providing statistically significant genetic correlations (*rg*) with either SGIT or UGIT and *rg* magnitude greater than 0.25 were considered. These traits were grouped into 11 manually annotated clusters. From each cluster we selected one trait (top-trait) providing the smallest p-value of genetic correlation among all pairwise correlations with SGIT and UGIT within this cluster and depicted this value on a heatmap. Genetic correlations not passing the significance threshold of p-value < 9.41e-07 are crossed out.

#### Polygenic risk score for SGIT and UGIT. Their role in disease/medical intervention prediction

We developed a model of PRS estimation using a training sample. For a testing sample, we calculated SGIT and UGIT PRSs using individual genotypes. Then we examined association between these PRSs and medical codes available for members of testing sample (see Supplementary Table ST10). We detected 92 ICD10 codes and 57 OPCS4 codes statistically significantly associated with at least one of the studied PRSs. SGIT PRS was associated with all of them. ICD10 codes associated with SGIT and/or UGIT PRSs related to a wide range of disorders, such as disorders of musculoskeletal, nervous, cardio-vascular, digestive and excretory systems, metabolic and skin disorders, diabetes, respiratory and neurological diseases (Supplementary Table ST10, Figure 6). The associated OPCS4 codes represented mostly surgical manipulations on joints and bones and diagnostic endoscopic examination of gastrointestinal and upper respiratory tract. Also, we observed SGIT PRS to be associated with all of the chronic pain phenotypes and these associations were characterized by the lowest p-values (Supplementary Table ST10). Unlike SGIT PRS, UGIT PRS provided only three significant associations, which had been detected for SGIT PRS as well: associations with chronic back and knee pain and with A52 OPCS4 code denoting therapeutic epidural injection, which is due to the association with chronic back pain (Supplementary Table ST10, Figure 6).

**Figure 6.**
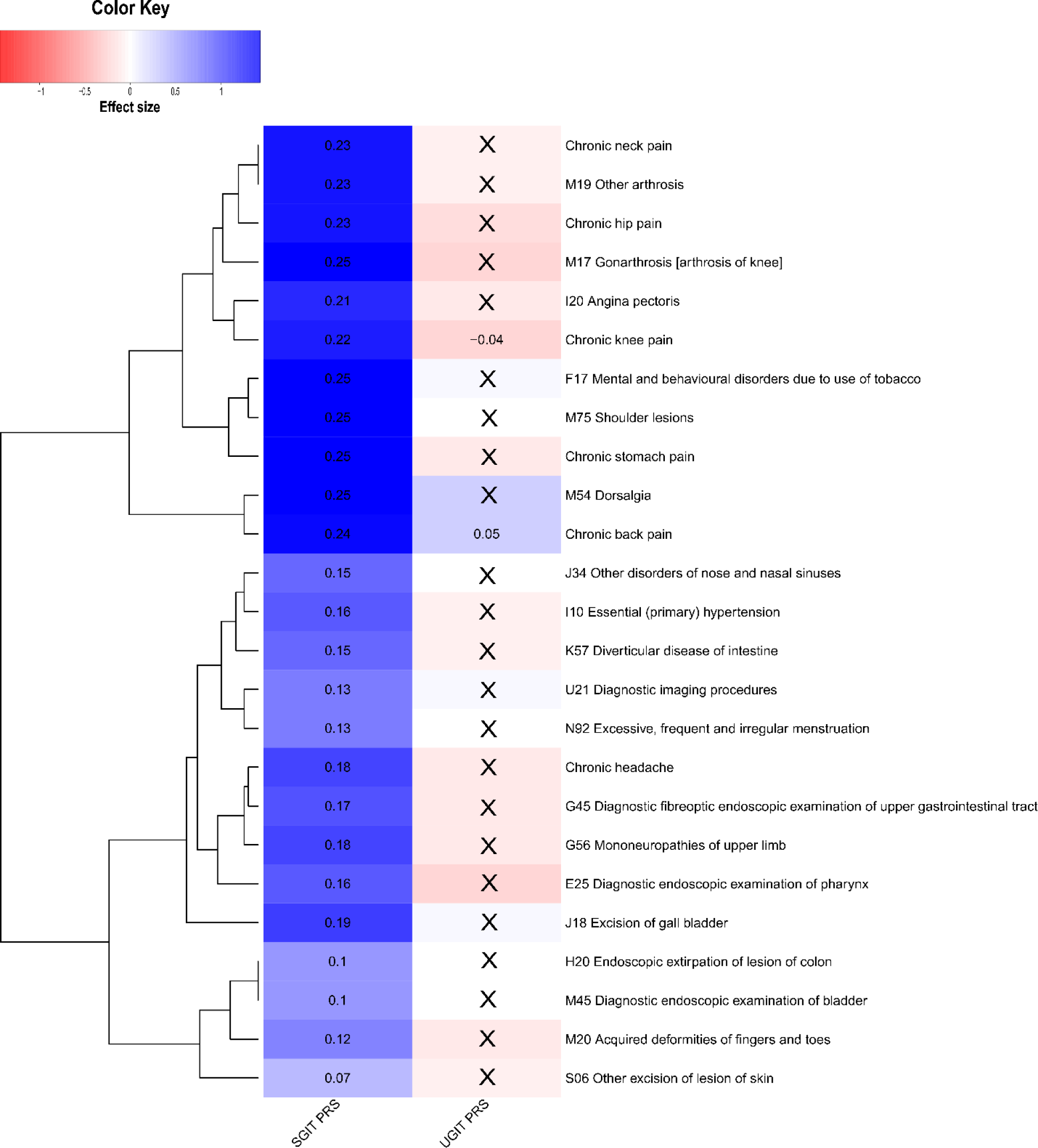
Associations of SGIT and UGIT PRSs with ICD10 and OPCS4 codes and chronic pain phenotypes. Only 92 ICD10 and 57 OPCS4 codes providing statistically significant associations with either CBP, SGIT or UGIT PRS were considered. Medical codes were grouped into 19 clusters. From each cluster we selected one code providing the smallest p-value of association with PRSs and depicted its effect sizes on a heatmap. Associations not passing the significance threshold of p-value < 8.25e-05 were crossed out.

### CBP subphenotyping among people with CBP

We investigated the potential of SGIT and UGIT binary-coded PRSs for CBP subphenotyping using only the 65,011 participants with CBP. For each individual and each of two traits, SGIT and UGIT, we formed two types of binary-coded PRSs defined by the lowest and highest deciles of risk (see “Polygenic risk score calculation” in Supplementary Methods section). Then we performed an analysis of associations between each of the four binary-coded PRSs (with two reflecting the lowest decile of the PRS for SGIT and UGIT, respectively, and the other two reflecting the highest decile- for each of SGIT and UGIT) and the ICD10/OPCS4 medical codes and identified 69 ICD10 and 35 OPCS4 codes statistically significantly associated with at least one of binary-coded PRSs (Supplementary Table ST11).

Being in the lowest decile of the SGIT PRS (low genetic predisposition to CBP through shared genetic background) provided a “unique” (not observed for other binary PRS traits) negative association with acute myocardial infarction (I21 ICD10 code, OR = 0.60, p-value = 1.09e-06). At the same time, the lowest decile of UGIT PRS (low genetic predisposition to CBP through the unshared genetic background) was uniquely positively associated with unspecified diabetes mellitus (E14 ICD10 code, OR = 1.64, p-value = 3.05e-05).

The highest decile of the SGIT PRS (high genetic predisposition to CBP through the shared genetic background) showed 102 statistically significant positive associations with the rest medical codes and 57 of them were associated only with this binary PRS trait. Among these 57 associated codes were those related to diagnostic endoscopic examination of joints and rheumatoid arthritis, surgical manipulation on muscles, joints and bones (such as repair of muscle, bone excision, replacement of joint etc.), conditions of the spine and nervous system, disorders of digestive, genitourinary and endocrine systems and neurological disorders. No specific association was found with the highest decile of the UGIT PRS, but differences in association pattern from other binary PRS traits were found. For example, the SGIT PRS was positively associated with gastrointestinal tract examination procedures (may be because these people are generally prone to pain, so they have unexplained abdominal pain as well or they may take NSAIDs), obesity, arthrosis of knee and hip, and knee surgery, while the UGIT PRS was negative associated with these traits.

Generally, the lowest decile of SGIT PRS and the highest decile of UGIT PRS manifested protective effects on medical states being diagnosed and treated, whereas the lowest decile of UGIT PRS and the highest decile of SGIT PRS had the opposite effect (Supplementary Table ST11).

## Discussion

In this work, we first applied the decomposition of genetic background of CBP into shared and unshared ones and showed that they differ in their functions. The shared genetic background is common to different chronic pain conditions while the unshared genetic background is related only to CBP. We built two traits, SGIT and UGIT, corresponding to the shared and unshared genetic background, respectively, and analyzed their properties.

We identified five loci associated with SGIT and one locus related to UGIT. Among them, only one was new – the locus on chromosome 17 associated with CBP through SGIT. We prioritized two genes (*CA10* and *LINC01982*) near it which are potentially involved in brain development, synapse formation and carcinogenesis, and found associations with gastroesophageal reflux, bone disease, multisite chronic pain and various known risk factors of chronic back pain (educational attainment, smoking, depression). The other three loci found associated with SGIT had previously been reported for GIP1 [16], as expected given our assumption that GIP1 is an approximation of the shared genetic background of the four chronic pain phenotypes studied before. In past work, we have prioritized 12 genes (*MIR7114*, *NOXA1*, *GRIN1*, *NSMF*, *FOXP2*, *BSN*, *AMIGO3*, *RBM6*, *FAM212A*, *RNF123*, *UBA7*, *ECM1*) in these three loci associated with SGIT, with six of them (*NOXA1*, *GRIN1*, *NSMF*, *FOXP2*, *BSN*, *AMIGO3*) being related to the nervous system (brain development and recovery, synapse plasticity, signal transduction, neuropathic and inflammatory pain). Some other genes were related to musculoskeletal (*MIR7114* and *ECM1,* functional in osteoarthritis and osteogenesis, respectively) and immune (*UBA7*) system processes.

In addition to five loci significantly associated with SGIT, there were four more loci suggestively associated (p-value < 8.3e-08) with SGIT (Supplementary Table ST12). Among these four loci there was a locus of *SLC39A8* gene reported in our previous study of GIP [16] and there was one new replicated under p-value < 8.3e-08 threshold locus on chromosome 13 with the lead SNP rs2587363, located in the *AL356295.1* gene (Supplementary Figure 6). We cannot consider this novel locus as statistically significant in the present study, however, it may nevertheless be a target for further investigation. This locus has been reported as associated with multisite chronic pain in females [37], major depressive disorder, osteoarthritis, post-traumatic stress disorder [38,39], and chronic fatigue syndrome [40]. Moreover, this locus contained polymorphism rs2587363 that was classified as pathogenic according to the FATHMM-XF (Supplementary Table ST13) and demonstrated a pleiotropic effect on the *OLFM4* gene expression (Supplementary Table ST14) in peripheral blood. This gene encodes olfactomedin 4, which is an antiapoptotic factor promoting tumor growth.

Additionally, we performed gene-based association analyses and identified five genes (*SLC39A8, FOXP2, MAML3, TSF20, GABRB2*) significantly associated with SGIT. As can be seen, *FOXP2* was observed in this study both in GWAS and in the gene-based analysis. The *SLC39A8* gene has been reported and replicated for GIP1 [16]. The product of the *SLC39A8* gene is a metal transporter with a role in manganese (Mn) homeostasis, and the missense rs13107325 in this gene is among the top pleiotropic SNPs identified in GWAS. Specifically, it has previously been associated with increased risk of osteoarthritis [41] and severe adolescent idiopathic scoliosis [42]. The gene is known to be mutated in congenital disorder of glycosylation, *SLC39A8*-CDG, with clinical features including osteopenia, broadened long bone epiphysis and joint hypermobility [43,44]. The dietary manganese deficiency is known to lead to bone and connective tissue disease in animals [45]. *MAML3* has been shown to contribute to several pathways significantly associated with CBP [46]. This gene enables transcription coactivator activity; it is involved in the Notch signaling pathway and positive regulation of transcription by RNA polymerase II. *GABRB2* encodes a GABA (gamma-aminobutyric acid) type A receptor beta subunit. The gene has a pivotal role in the central nervous system and associates with various neuropsychiatric disorders [47]. *TCF20* encodes a transcription factor that recognizes the platelet-derived growth factor-responsive element in the matrix metalloproteinase 3 promoter. The encoded protein is thought to be a transcriptional coactivator, enhancing the activity of transcription factors such as JUN and SP1. Mutations in this gene are associated with autism spectrum disorders (according to NCBI-RefSeq https://www.ncbi.nlm.nih.gov/gene?Db=gene&Cmd=DetailsSearch&Term=6942). Recently, it has been shown that *TCF20* is essential for neurogenesis during embryonic brain development in mouse. *TCF20* dysfunction leads to deficits in neurogenesis, which further results in the development of autism spectrum disorders [48].

Recently, more has been paid to the study of the role of rare genetic variants in the control of different traits. In the current study, we also tested the association of SGIT with exome sequencing data from UK Biobank (Supplementary Methods) and performed gene-based association analysis, which identified the *SLC13A1* gene (Supplementary Tables ST15a, b). The effect of this gene was explained by its loss of function (LoF) and missense variants. The *SLC13A1* gene has been previously detected as associated with back pain-related traits due to LoF variants [14,49]. This gene encodes a protein that functions as a high-affinity sodium-dependent sulfate transmembrane transporter [50]. A deficiency of the SLC13A1 protein is associated with a reduced blood sulfate level, which plays a key role in the underlying processes leading to painful intervertebral disc disorders [14]. The association of a gene with disease through LoF variants presents several advantages as a drug target because it provides a clear functional link, target specificity, potential for gene therapy, predictive value, and personalized medicine opportunities. Until now, this gene has been considered as a potential target for treatment against back pain. In the light of our results, *SLC13A1* can be considered as a potential target for treatment against chronic pain irrespective of location.

For UGIT, the *SPOCK2* and *CHST3* genes were detected using both gene-based and gene prioritization analysis. The association of *SPOCK2* with back pain has been described previously [15]. This gene is located close to carbohydrate sulfotransferase 3 (*CHST3*) which codes for an enzyme that catalyzes proteoglycan sulfating and has been identified as a susceptibility gene for lumbar disc degeneration [51]. The association of *CHST3* with back pain-related phenotypes has also been described [14]. This gene can be considered as most probably causal for back pain in this region, since cumulative evidence L2G scores for *CHST3* and *SPOCK2* are 0.81 and 0.23, respectively, https://genetics.opentargets.org/study-locus/NEALE2_6159_4/10_72001257_A_G, [52].

Information on genes associated with SGIT is potentially useful in terms of discovering biomarkers and developing multi-target drugs against chronic pain at several anatomic sites at once, because these genes are most likely involved in the development of chronic pain in general. In contrast, genes related to the unshared genetic background of CBP may be more pertinent to the development of chronic pain at back precisely, probably by affecting chondrogenesis or degenerative processes in spine. So UGIT associated genes may be of interest for the development of drugs for treatment of chronic pain specifically in the back.

Based on enrichment analysis and functional annotation of associated loci, the shared genetic background common to different chronic pain conditions contributes to these phenotypes in general through processes in the central nervous and neurosecretory systems. SGIT genetic correlations provided evidence of genetically predisposed psychosocial aspects of CBP. The SNP-based heritability for UGIT was much smaller than that of SGIT, resulting in much less statistical power for all analyses. However, we still detected some significant results. A few findings from functional annotation demonstrated that UGIT was inversely associated with knee pain and headache and positively associated with back pain and medical treatments for back pain such as epidural injection. UGIT also tended to be negatively correlated with respiratory illnesses, musculoskeletal disorders and injuries, demographic parameters and psychometric disorders, whereas SGIT is positively associated with all of these traits. These findings taken together support that the unshared genetic background of CBP may determine the development of pain in the back specifically, whereas the shared genetic background controls non-specific pain manifestations and processes accompanying different pain conditions.

Genetic correlation analysis and PRS analysis are two ways to investigate the influence of genetic background of CBP. Although the lists of the traits included in these analyses overlap, there are many traits presented only in one of them. Thus, genetic correlation analysis adds information on association with self-reported and anthropometric traits (e.g., UGIT is significantly correlated with height), and PRS analysis provides more information on medical diagnoses and interventions (for instance, both SGIT and UGIT PRSs are positively associated with therapeutic epidural injection). Findings from the two methods complement each other. For example, genetic correlation analysis of SGIT reveals genetic predisposition to greater alcohol intake and smoking, and in PRS analysis, we observe its association with mental disorders due to alcohol intake and smoking. Similarly, SGIT analysis shows positive genetic correlation with toothache, and PRS analysis highlights association with tooth removal. For group of traits included in both analyses (such as lower respiratory and urinary system disorders, anxiety and depression, intestine and cardio-vascular diseases, sleep and eye disorders, arthritis, spine and knee problems, etc.) results suggest positive association with SGIT in both methods, but they should not be interpreted in the same way. While genetic correlation analysis provides information on pleiotropic effects or linkage in genome [53], PRS analysis reveals associations between the genetic background of CBP and phenotypic traits that can be explained not only by linkage or pleiotropic effects of genes, but also by causal effects of CBP on other traits. Comparing results of genetic correlation analysis with those obtained from PRS analysis can tell us more about probable reasons of observed association between two phenotypes [54]. For instance, when we observe modest genetic correlation between traits along with high magnitude of regression coefficient from PRS analysis, this may indicate the role of causal or confounding effects, rather than effects of genetic pleiotropy or linkage. Trait pairs with such a pattern may be an interesting subject for further studies utilizing Mendelian randomization. For example, we observed this pattern for SGIT and several traits, including obesity (*rg* with BMI was 0.31, beta from PRS analysis for obesity was 0.25), diabetes (*rg* was approximately 0.31 for set of related traits, PRS beta = 0.21), and migraine (*rg* = 0.37, PRS beta = 0.22). Causal effects of increased BMI on back pain, and back pain on type II diabetes, have already been reported in our studies [55,56], while the association with migraine is yet to be examined. In contrast, if the magnitude of pairwise genetic correlation coefficient is quite high it is likely that the phenotypic association between two traits is explained by similarity of their genetic background and not by causality. For pairs of traits with these characteristics, information on the similarity of underlying genetic mechanisms can be valuable for development or repurposing of medications affecting both traits via the same pathway. From this prospective, further in-depth studies of the high genetic correlation between irritable bowel syndrome and SGIT (*rg* = 0.78) might be worthwhile.

The results of genetic analysis of SGIT and UGIT allowed us to estimate their polygenic risk scores for participants with CBP and show that they are associated with different ICD10 and OPCS4 coded conditions. We introduced the binary-coded PRSs for SGIT and UGIT to characterize low and high genetic risk of CBP related to its shared and unshared genetic background, respectively. We showed that these binary-coded PRSs can be potentially helpful for subphenotyping of individuals with CBP, because participants with low or high genetic risk of CBP defined by the shared or unshared genetic background are characterized with different patterns of association with medical interventions and disorders. This can be crucial for forecasting pain trajectories and choosing an adequate treatment. Low genetic liability to CBP explained by shared genetic background turned out to be protective against diagnosis of acute myocardial infarction (OR = 0.60, p-value = 1.09e-06), but low genetic risk of CBP mediated through its unshared genetic background was associated with higher predisposition to unspecified diabetes mellitus (OR = 1.64, p-value = 3.05e-05). High PRS of SGIT significantly increased the risk of diverse diseases of musculoskeletal, nervous, digestive, genitourinary and endocrine systems (ORs varying from 1.28 to 2.08). Genetic liability to CBP through shared genetic background also showed positive association with gastrointestinal tract examination procedures, this may be because people are generally prone to pain, so they have unexplained abdominal pain as well or they may take non-steroidal anti-inflammatory drugs, which have side effects on gastrointestinal system. All these results should be interpreted with caution, since negative/positive association between PRS and medical code does not necessarily mean protective/detrimental effect on disease development itself but on the disease being diagnosed. For instance, low OR of myocardial infarction among people with low SGIT PRS could potentially indicate that their low genetic liability to pain regardless of its site (including chest pain) can make myocardial infarction much harder to detect. Such a phenomenon was observed for diabetics, who often do not have chest pain, so their myocardial infarctions do not get diagnosed [57,58].

Our study has some limitations and restrictions. First, the discovery analyses were performed in Northern Europeans, so the obtained linear combination coefficients for SGIT and UGIT cannot strictly be applied to individuals of African and Asian ancestry, included in replication sample. Secondly, both discovery and replication sample were based on UK Biobank participants, which means that other cohorts are needed to perform independent replication of the results. Thirdly, here we refer to UGIT as pertaining to unshared genetic background specific to CBP, however, the SHAHER approach does not necessarily accurately divide the genetic background of the traits, so UGIT may have genetic correlations with some of the other chronic pain traits but not with all of them. Finally, we had limited statistical power in UGIT functional analyses, which resulted in few significant findings. This indicates that larger sample size may be helpful in further studies.

Overall, the current work demonstrates that genetic background of CBP can be split into shared between different pain conditions and specific for CBP background. The former is likely to maintain pain mechanisms and manifestations in general through the central nervous and immune systems, functioning of joints, general processes of bone formation and remodeling, while the latter is responsible for processes specific to back pain. Polygenic risk scores separately accounting for shared and unshared genetic background of CBP identify subphenotypes characterized by different genetic predisposition to a range of medical conditions making them possible tools for prognosis in the healthcare system. Twenty genes prioritized for CBP may provide further opportunities for advances in chronic pain management, such as the discovery of new biomarkers and drug targets for chronic pain regardless of its site and drug targets that are specific to CBP.

## Materials and Methods

### Data description

We used the UK Biobank GWAS data (Ntotal = 456,000) relating to four chronic musculoskeletal pain traits (pain in the back, neck, hip, knee) from our recent work (see [16] for the detailed description of the phenotype definition, sample size and sample characteristics). We obtained the remaining two chronic pain phenotypes (stomach pain and headache) from the same UK Biobank [59] sample and conducted GWAS according to the protocol we have described previously (Supplementary Methods, [16]). For each pain trait we split the whole sample of UK Biobank participants into a discovery (N = 265,000, European ancestry individuals) and a replication sample (three samples of African, N = 7,541, South Asian, N = 9,208, and European descent individuals, N = 174,831). Details of the samples (size, sex and age structure, pain type prevalence and BMI distribution) are available in Supplementary Table ST1. Data were uploaded to the GWAS-Map database [29,30] for quality control, unification and further functional analysis of GWAS summary statistics. The same genotype and phenotype data were used for making and testing PRS models.

### Decomposition of genetic background of chronic back pain

To decompose the genetic background of CBP we implemented the SHAHER framework [25]. This approach is based on the concept that genetic background of each of the genetically related traits can be decomposed into two parts reflecting common for all traits (shared genetic background) and specific for particular trait background. To identify shared and unshared (or specific) background of CBP, SHAHER composes two new traits: SGIT, which condenses the shared genetic background, and UGIT, which is controlled by specific for CBP genetic background. Both SGIT and UGIT are considered as linear combinations of the original traits with specific coefficients. The coefficients for SGIT are calculated by maximizing the proportion of CBP shared genetic background in SGIT genetic background. To calculate the coefficients for UGIT, we estimated the residual genetic background of CBP after adjustment for SGIT. Using these coefficients and the GWAS summary statistics for all original traits, the GWAS summary statistics for SGIT and UGIT of CBP were calculated.

To estimate the SGIT coefficients we utilized the heritability estimates, phenotypic correlation matrices, and genetic correlation matrices (see details in Supplementary Methods) assessed beforehand for the original pain traits in the discovery sample. Further, we used the coefficients obtained using discovery sample in SHAHER analysis of all replication samples (Figure 1).

### Meta-analysis of GWAS summary statistics using METAL

For both SGIT and UGIT we performed two GWAS meta-analyses: (i) a meta-analysis comprising all replication samples (Replication meta-analysis, Ntotal = 191,580); and (ii) a meta-analysis of two European samples (European meta-analysis, Ntotal = 439,831). The first meta-analysis served as a replication set for the GWAS associations observed in the discovery set, while the second one was utilized for further functional annotation. In a gene-based analysis we utilized European meta-analysis for additional replication of the signals from the discovery sample. All the meta-analyses were conducted using an inverse-variance-weighted approach and fixed-effects model implemented in METAL software, version 2011-03-25 [60].

### Gene identification

#### Loci identification and replication

In order to identify loci statistically significantly associated with either SGIT or UGIT we carried out the conditional and joint analysis using GCTA-COJO software (version 1.90.0beta [61]) on the discovery sample. We chose the following settings: minor allele frequency (MAF) not less than 2e-04; stepwise model selection procedure to select independently associated SNPs; significance threshold (applied to statistics after correction for genomic control using the LD Score regression intercept) p-value = 5e-08/6 = 8.3e-09, where the denominator corresponds to the number of analyzed traits (SGIT and UGIT from the current work, as well as four genetically independent phenotypes [GIP1-4] from our recent study [16] which were used for comparison); and ±5,000 kb window for linkage disequilibrium assessment. A linkage disequilibrium matrix was computed using PLINK version 1.9 (https://www.cog-genomics.org/plink/1.9/) using 100,000 randomly selected individuals from the discovery set. Loci associated with either SGIT or UGIT were defined as genomic regions of ±250 kb from the lead SNPs identified in COJO analysis.

For both SGIT-associated and UGIT-associated loci we performed a two-step replication procedure. First, we checked whether associations were observed in previous pain studies. We compared SGIT-associated regions with those reported for the first genetically independent phenotype (GIP1) [16] and collated UGIT-associated signal with results from an earlier back pain GWAS [15]. Loci demonstrating the same direction of the effect in the current and previous studies were considered as *known*. Second, we focused on *new* signals (not reported previously) and examined them in the replication meta-analysis. We assumed new signals to be *replicated* if two criteria were met: 1) association in the replication meta-analysis was statistically significant after Bonferroni correction [62] for multiple testing (p-value < 0.05/*n*, where *n* is a number of new loci associated with either SGIT or UGIT); and 2) direction of the effect was the same in the discovery sample and Replication meta-analysis.

#### Gene prioritization in associated loci

We prioritized genes in the replicated locus using a protocol we have described previously in our work on GIP [16]. For gene prioritization we applied a series of methods including (1) literature-based annotation of all genes within the locus using various databases; (2) prediction of SNP effects in the locus utilizing Ensembl Variant Effect Predictor (VEP) [31], FATHMM-XF [32] and FATHMM-INDEL [33] tools; (3) gene prioritization embedded into Data-driven Expression Prioritized Integration for Complex Traits (DEPICT) software [34], and Functional Mapping and Annotation tool (FUMA) [35]; (4) analysis of pleiotropic effects of the replicated loci on gene expression in various tissue types (see Supplementary Table ST16) using an instrument combining Mendelian randomization (Summary data-based Mendelian Randomization, SMR) with heterogeneity testing (Heterogeneity in Dependent Instruments, HEIDI) [36]. Genes provided more evidence for prioritization were considered as more likely to be causal. Details are available in the Supplementary Methods.

#### Gene-based association analysis

We conducted a gene-based association analysis of SGIT and UGIT utilizing GWAS summary statistics (z-scores and effect sizes) from three datasets: the discovery sample (N = 265,000), the European replication sample (N = 174,831), and data from the European meta-analysis (Ntotal = 439,831). We preliminarily calculated the matrices of correlations between genotypes of all SNPs within a gene using individual genotypes of non-relatives from the entire European sample of UK Biobank participants (N = 315,599) and the PLINK software v2.00a3.7LM (https://www.cog-genomics.org/plink/2.0/) with options –maf 5e-05 --geno 0.02. For replication, we restricted to the European sample because discovery and replication gene-based analyses had to be performed using the same matrix of genotype correlations, which is ancestry-specific. In the discovery sample we set the Bonferroni adjusted significance level [62] for the total number of genes (20,000) at 2.5e-06. We defined two criteria for replication of a gene-based signal: 1) the p-value in European replication sample had to be less than 0.05/*k*, where *k* corresponds to the number of statistically significant signals in the discovery sample; and 2) the p-value in European meta-analysis had to be less then the p-value in the discovery sample. See Supplementary Methods for more details on genomic regions studied, methods of gene-based and conditional analyses.

### Analysis of CBP genetic architecture

#### Gene set and tissue/cell type enrichment

We conducted DEPICT tissue or cell type enrichment analyses for GWAS loci associated with SGIT or UGIT in European meta-analysis. We used DEPICT software, version 1.1 rel194 [34], with default settings. For each of the traits we analyzed two SNP lists: a set of variants associated with the trait under p-value < 2.5e-08 significance threshold, and a set of SNPs with p-value < 5e-06. The significance thresholds corresponds to those recommended by program developers with correction for multiple testing for two traits applied. Variants were selected using GCTA-COJO instrument with settings described above in ‘Loci identification and replication’ section.

For all genes identified for SGIT we ran the gene set enrichment and tissue specificity analyses utilizing FUMA function GENE2FUNC. As the input data we used: 1) all genes prioritized for SGIT in the GWAS loci (both from the new replicated locus and from the loci previously replicated for GIP1); 2) all genes found in gene-based analysis and replicated in European meta-analysis. We analyzed SGIT-associated genes only, because there were not enough genes identified for UGIT to perform the analysis. The GTEx v8 dataset for 30 general tissues was used for tissue specificity analysis. A set of the input genes was tested against each of the sets of differentially expressed genes (DEG) using a hypergeometric test and Bonferroni multiple testing correction. Statistical significance was determined at an adjusted p-value < 0.05. We estimated the overrepresentation of the identified genes in the gene sets of the GWAS catalog. Hypergeometric tests were performed to check if the genes of interest were overrepresented in any of the pre-defined gene sets. In this type of analysis, we used FDR (Benjamini – Hochberg method) for multiple testing correction as was recommended by FUMA. Statistical significance was determined at a q-value < 0.05.

#### Genetic correlation between SGIT, UGIT and complex traits

We assessed genetic correlations of SGIT and UGIT with 730 complex human traits from the GWAS-Map database using LD Score regression tool embedded in the platform. The detailed protocol of selection of 730 complex traits included in the analysis and the full list of them were provided in a recent study by Timmers et al. [63]. For further interpretation we restricted our analysis to 322 complex traits which had statistically significant genetic correlations with SGIT and/or UGIT with magnitude greater than 0.25 (p-value < 3.42e-5 = 0.05/(730*2), where 730 represents the total number of complex traits and 2 refers to the number of traits). To simplify the perception and visualization of the results of genetic correlation analysis we performed a hierarchal clustering of 322 complex traits. This was done utilizing the ward.D2 method from the standard hclust() R function on a squared matrix of genetic correlations transformed to the Euclidian distances (as.dist() R function) as described previously by Tsepilov et al. [16]. We set the arbitrary threshold of h = 1.8 to cut the dendrogram of hierarchical clustering by height and obtained 11 clusters, each of which was then annotated manually. The full list of 322 traits grouped by clusters is provided in Supplementary Table ST9.

Heatmap visualization of the genetic correlations was made using the heatmap.2() function from the gplots R package, version 3.1.1. From each cluster, we depicted one trait having the lowest p-value of genetic correlation among all pairwise correlations with SGIT and UGIT within the cluster. The threshold for statistical significance was p-value > 9.41e-07 = 0.05/(51,681 + 730*2), where 51,681 is the number of unique genetic correlation coefficients in the squared matrix for 322 complex traits, and 730*2 is the total number of genetic correlations between SGIT, UGIT and 730 complex traits.

#### Polygenic risk scores of SGIT and UGIT. Their role in disease/medical intervention prediction

In addition to other functional analyses, we examined the prognostic power of SGIT and UGIT polygenic risk scores (PRS) and assessed their role in disease/medical intervention prediction. We calculated the PRSs for SGIT and UGIT utilizing data from the entire European sample (discovery and European replication samples, Ntotal = 439,762) using a three-step algorithm. We used the SBayesR method [64] and the protocol from our recent work on CBP PRS [24], with some modifications. We divided the entire European sample into a training set (discovery sample, N = 265,000), a validation set (subsample from the European replication sample, N = 30,000), and a test set (the other 144,831 Northern Europeans from the European replication sample). The first set was used to develop PRS models, the second one was used to validate these models in individual-level data and select the optimal models for SGIT and UGIT, and the last dataset was used to assess quality metrics of the optimal models. At the final step, PRS values were calculated for all individuals from entire European sample using optimal models. More details on PRS calculation available in Supplementary Methods.

When the PRS for SGIT and UGIT were estimated for the entire European sample of 439,831, we normalized the PRS (the variance was taken to 1) and used them in a number of generalized linear models (GLMs) considering PRS as a predictor for a particular trait. List of the traits and corresponding phenotypic data were taken from medical histories and questionnaires obtained from UK Biobank participants of European descent from the test dataset mentioned in the paragraph above, using non-relatives only (N = 120,200). Non-relatives were defined by UK Biobank data-field 22021 (for more details see https://biobank.ndph.ox.ac.uk/showcase/coding.cgi?id=682).

Medical codes were combined to the second level, meaning that participants with a specific medical code of the second level and/or codes of a lower level (the third and/or the fourth) nested under the second level code were all considered to be cases of the second level medical code (for more information on ICD10 and OPCS4 coding in the UK Biobank see https://biobank.ndph.ox.ac.uk/ukb/field.cgi?id=41202 and https://biobank.ndph.ox.ac.uk/ukb/field.cgi?id=41200). Only ICD10 codes from chapters I-XVII were analyzed, OPCS4 codes from chapters X, Y, Z were excluded, and then the remaining codes were filtered by prevalence (> 0.5% and < 99.5%). The final list of the traits (see Supplementary Table ST10) comprised the six self-reported chronic pain phenotypes (questionnaire-based) considered in the study, 165 ICD10 and 132 OPCS4 medical codes served as proxies for the corresponding diseases/medical interventions. To perform GLM-analyses we used a logistic regression from a standard glm() function in R and included sex, age, batch number and the first ten genetic principal components (PC1-10) provided by UK Biobank as covariates in addition to the PRS predictor. The general formula is the following: medical code ∼ age + sex + batch + PC1 + … + PC10 + PRS. Finally, we filtered out the GLM results not passing the significance threshold of p-value < 8.25e-05 = 0.05/(2*(165 + 132 + 6)), where 165 and 132 are the numbers of ICD10 and OPCS4 codes, respectively, 6 is a number of pain traits and 2 corresponds to the number of PRSs. For further heatmap visualization of the significant GLM-results we performed a hierarchical clustering of medical codes based on their phenotypic correlation matrix as described above in the genetic correlation analysis. We set a h = 1.25 threshold for cutting the hierarchical clustering dendrogram by height and defined 19 clusters (see Supplementary Table ST10 for more details). Similarly, for the genetic correlation analysis we selected one medical code from each cluster, which provided the lowest p-value of association with SGIT and UGIT polygenic risk scores. Alongside medical codes, representing particular clusters, we added six pain traits to the heatmap plot. The visualization was made as described in the genetic correlation analysis section.

### CBP subphenotyping among people with CBP

To investigate the potential of SGIT and UGIT PRSs for back pain subphenotyping, we analyzed only CBP cases from the entire European sample, focusing on non-relatives only (N = 65,011). The decision to switch to the entire European sample instead of working with the test sample only was motivated by the limited statistical power of the test sample. We normalized SGIT and UGIT PRS values and recoded each of the vectors of normalized PRSs as binary traits in two different ways. First, we coded the normalized PRS values as 1 if a participant with back pain was attributed to the lowest decile of the normalized PRS vector for SGIT or UGIT, respectively. Normalized PRS values falling out of this range were coded as 0. By such a coding (“yes/no in the lowest decile of the normalized PRS values for SGIT”; “yes/no in the lowest decile of the normalized PRS values for UGIT”) we highlighted individuals with the lowest genetic predisposition to CBP through SGIT and UGIT. Then, we coded as 1 the normalized PRS estimates referring to the highest decile of the SGIT or UGIT PRS distribution, respectively, and coded them as 0 otherwise. In this binary coding (“yes/no in the highest decile of the normalized PRS values for SGIT”; “yes/no in the highest decile of the normalized PRS values for UGIT”) we marked individuals with the highest genetic predisposition to CBP through SGIT and UGIT. For each of these binary traits we performed GLM analyses using the model described above. As a dependent variable, we considered ICD10 and OPCS4 codes described above. In this sample of 65,011 non-related participants with CBP we selected 199 ICD10 and 165 OPCS4 codes with prevalence > 0.5% and < 99.5%. The significance threshold for this analysis was set at p = 3.43e-05 = 0.05/(4*(199 + 165)), where 4 is the number of binary PRS traits, 199 is the number of ICD10 codes, and 165 is the number of OPCS4 codes.

## Data and code availability

Full project code is available at https://github.com/ElizavetaElgaeva/BP-SH_project_code.

GWAS and EWAS summary statistics for all pain traits along with SGIT and UGIT PRS model weights can be found here (the link will be added later).

## Supporting information

Supplementary Results

## Data Availability

The project code used for performing analysis is currently in open access. All the summary-level data produced in the course of this work will be available for download via the link to our site after publication of the article.

## Acknowledgements

This study was conducted using the UK Biobank Resource (project #18219 and #59345). The work of ANT is an output of a research project implemented as part of the Research Program at the MSU Institute for Artificial Intelligence. FMKW was supported by Versus Arthritis, grant #22467. We thank Dr. Alexandra S. Shadrina for providing template for Figure 1 and Dr. Sodbo Z. Sharapov for advices on study design.

## Funding

IVZ, YAT, GRS, and AVK were supported by the budget project of the Institute of Cytology and Genetics FWNR-2022-0020. The work of TIA and EEE was supported by the Russian Science Foundation (grant #22-15-20037) and Government of the Novosibirsk region.

## Authors’ contributions

EEE participated in conceptualization, data curation, formal analysis, investigation, project administration, software, supervision, validation, visualization, and original draft preparation. IVZ contributed to formal analysis, investigation, software, validation, visualization, and original draft preparation. AVN took part in data curation, formal analysis, software, methodology, validation, visualization, and original draft preparation. DAV carried out formal analysis, software and investigation. EST participated in data curation, formal analysis and software. ANT contributed in formal analysis and software. AVK provided data curation, formal analysis, resources and software. GRS contributed in methodology and software. MBF, FMKW, PS and YSA carried out data curation. TIA, YAT, YSA, FMKW, and PS participated in conceptualization, investigation, project administration, and supervision. TIA contributed to visualization. YAT provided funding acquisition. All co-authors discussed the results and contributed to review and editing while preparing the final version of the manuscript. All authors have read and agreed to the published version of the manuscript.

## Conflict of interest

YSA is a cofounder and a co-owner of PolyOmica and PolyKnomics, private organizations providing services, research, and development in the field of computational and statistical genomics.

## Supplementary note

Supplementary Methods

Extended description of the Materials and Methods section.

Supplementary Results

Detailed description of the results of gene prioritization.

Supplementary Figure 1

Manhattan plot for SGIT in discovery sample.

Supplementary Figure 2

Quantile-quantile (QQ) plot for SGIT in discovery sample.

Supplementary Figure 3

Manhattan plot for UGIT in discovery sample.

Supplementary Figure 4

Quantile-quantile (QQ) plot for UGIT in discovery sample.

Supplementary Figure 5

Gene expression patterns of 23 genes identified for SGIT.

Supplementary Figure 6

Regional association plot for rs2587363 locus associated with SGIT under suggestive significance threshold p-value < 8.3e-08.

Supplementary Table ST1

Description of the sample.

**Supplementary Table ST2** Characteristics of pain traits.

**Supplementary Table ST3**

Top loci associated with SGIT and UGIT at a study-level threshold of statistical significance (p- value < 8.3e-09).

Supplementary Table ST4

Results of SNP effect prediction using Variant Effect Predictor and FATMM tools.

Supplementary Table ST5

Results of gene prioritization using DEPICT for SNPs associated with SGIT in European meta- analysis.

Supplementary Table ST6

Results of FUMA gene prioritization for SNPs associated with SGIT in European meta-analysis at p-value < 2.5e-08 and p-value < 5e-06.

Supplementary Table ST7

Results of gene-based and conditional analyses.

Supplementary Table ST8

Results of DEPICT analysis for SNPs associated with SGIT in European meta-analysis.

Supplementary Table ST9

Analysis of genetic correlations.

Supplementary Table ST10

Results of PRS analysis in the test sample, non-relatives only.

Supplementary Table ST11

Results of binary-coded PRS traits analysis in participants with back pain.

Supplementary Table ST12

Top loci associated with SGIT and UGIT under suggestive significance threshold p-value < 8.3e- 08.

Supplementary Table ST13

Results of SNP effect prediction using FATHMM-XF for the locus tagged by rs2587363.

Supplementary Table ST14

Results of SMR-HEIDI analysis of pleiotropic effects on gene expression for the locus tagged by rs2587363.

Supplementary Table ST15

Results of gene-based association analysis in exome data.

Supplementary Table ST16

List of tissues used for SMR-HEIDI analysis of pleiotropic effects on gene expression.

## Notes

### Author Declarations

All the source data were prodused prior to initiation of the current study and were provided by UK Biobank under projects #18219 and #59345. UK Biobank publically provide anonymized data under special request https://www.ukbiobank.ac.uk/enable-your-research/apply-for-access.

