## Supplementary Methods for "Decomposing the genetic background of chronic back pain"

### Pain phenotype definition

All study participants were asked about "Pain type(s) experienced in the last months" with following clarification whether the pain at specific location had been lasting more than 3 months. Individuals reported pain at the particular anatomic site present for more than 3 months were considered as cases of chronic pain at this specific location and were attributed to controls otherwise. Those who preferred not to answer or reported pain all over the body for more than 3 months were excluded from the study. Every UK Biobank participant has signed the voluntary informed consent.

### Sample size and power calculation

To obtain 80% statistical power, we split the entire European sample into the discovery and European replication samples as it was done in the same UK Biobank sample in our previous work [1]. The rationale for the proportion used for splitting the entire European sample into discovery and replication with corresponding power calculation are provided in [1].

### Estimation of heritability, genetic and phenotypic correlation matrices for pain traits

We estimated heritability, phenotypic correlations, and genetic correlations of the pain traits utilizing data from the discovery sample and tools embedded into the GWAS-Map platform. We assessed the SNP-based heritability and genetic correlation matrix using LD Score regression [2]. Pairwise phenotypic correlation coefficients were calculated according to the approach described by Stephens et al. [3]. The heatmap visualization of the correlation matrices was performed with the corrplot() function of the R package, version 0.92 [4].

### Gene prioritization in associated loci

All the gene prioritization analyses were performed utilizing GWAS summary statistics derived from European meta-analysis in order to obtain greater statistical power. We used several instruments for gene prioritization: literature-based annotation of the genes within the locus, prediction of SNP effects with VEP and FATMM, gene prioritization via DEPICT and FUMA, and analysis of pleotropic effects on gene expression using SMR-HEIDI (colocalization analysis).

Gene provided more pieces of evidence for prioritization by different methods was considered as more likely to be causal.

#### *Literature annotation of the genes within the loci*

We identified genes located in the replicated locus using the regional association plots obtained with the LocusZoom version 0.14 online tool (https://my.locuszoom.org/) [5]. For each of these genes we performed a literature-based annotation utilizing the following databases: GeneCards (https://www.genecards.org/), OMIM (https://www.omim.org/), Google Scholar, NCBI Gene (https://www.ncbi.nlm.nih.gov/gene) and PubMed.

#### *Prediction of SNP effects with VEP and FATHMM tools*

We performed an effect prediction analysis for replicated locus. Besides the lead SNP of this locus, identified by COJO, we also included polymorphisms and indels in high linkage disequilibrium ($r^2 > 0.8$) with it. To assess the linkage disequilibrium (LD) we applied PLINK software [6], version 1.9, --show-tags option, on genotype data from the 1000 Genomes project (phase 3, version 5, European ancestry individuals, N = 503). Moreover, we included SNPs from the replicated locus, having p-value ≤ T, where T was defined using the following equation: $\log10(T) = \log10(P_{min}) + 1$, and $P_{min}$ – is a p-value of the lead SNP in the region. By doing this, we aimed to extend the SNP list and account for those GWAS variants that could be potentially missed during the analysis, since the UK Biobank data were imputed using HRC, not the 1000 Genomes panel used for LD calculation. For all selected SNPs we performed a functional annotation utilizing Ensembl Variant Effect Predictor (VEP) [7], FATHMM-XF [8] and FATHMM-INDEL [9] tools. Effect prediction made by FATHMM was carried out based on a scale from 0 to 1, where values greater than 0.5 indicate deleterious mutations and values below the 0.5 threshold correspond to potentially beneficial or neutral variant substitution effects.

#### *Gene prioritization using DEPICT and FUMA*

For gene prioritization genomic regions we utilized the Data-driven Expression Prioritized Integration for Complex Traits (DEPICT) software, version 1.1 rel194 [10]. We used the default parameters (https://data.broadinstitute.org/mpg/depict/) and performed the analysis for SGIT and UGIT. We used four sets of SNPs as the input data: two sets under two significance thresholds (p-value < 2.5e-08 and p-value < 5e-06) both for SGIT and UGIT. The SNPs were selected from European meta-analysis data using COJO as it was described in loci identification part of the

‘Materials and Methods’ section in the main text. The use of two significance thresholds (5e-08 and 1e-05) is recommended by the DEPICT developers. Here we corrected both of these thresholds using Bonferroni correction for multiple testing [11] by dividing them by 2 to account for two traits. The MHC region was excluded from the DEPICT analysis. While interpreting the results a false discovery rate correction [12] FDR < 0.05 was applied.

Additionally, we performed gene prioritization with FUMA [13] using the SNP2GENE function (default settings). The significance threshold for FUMA analysis was defined as p-value < 0.05 after Bonferroni correction [11].

To prioritize genes in the single replicated locus associated with SGIT we extracted the information about this region from both DEPICT and FUMA results. Results for other genomic regions associated with SGIT and UGIT were used for general characterization, gene-set enrichment and tissue enrichment analyses.

#### *Colocalization analysis using SMR-HEIDI*

We examined the pleiotropic effects of the replicated locus on gene expression in various tissue types (see the list of analyzed tissues in Supplementary Table ST16) using a method integrating Mendelian randomization (Summary data-based Mendelian Randomization, SMR) and heterogeneity testing (Heterogeneity in Dependent Instruments, HEIDI) [14]. GWAS data on gene expression were obtained from Westra Blood eQTL (peripheral blood, https://yanglab.westlake.edu.cn/software/smr/#eQTLsummarydata) [15], Genotype-Tissue Expression (GTEx) version 7 (blood, musculoskeletal and nerve tissues, https://gtexportal.org) [16] and CEDAR projects (peripheral blood, http://cedar-web.giga.ulg.ac.be/) [17]. The analyses were performed in a direction from the genetic impact trait to gene expression, so they should be interpreted as colocalization analyses, not as testing for causal relationship. We set the significance threshold at p-value < 0.05/144 = 3.47e-04 for SMR analysis, where 144 corresponds to the total number of statistical tests, accounting for all gene expression profiles in different tissues available for analyzed SNPs. For the HEIDI test, p-value < 0.01 indicated the rejection of the null hypothesis regarding pleiotropy/colocalization.

### Gene-based association analysis

For gene-based association analysis, GWAS summary statistics (z-scores and effect sizes) for each variant, and the matrices of correlations between genotypes of all variants within a gene were calculated.

#### *Matrices of genotype correlations*

Correlation between every pair of variants within a gene was estimated using 315,599 unrelated white UK Biobank participants. PLINK v2.00a3.7LM (https://www.cog-genomics.org/plink/2.0/) with options –maf 5e-05 --geno 0.02 was used to select SNPs. SNP correlations were estimated using LDstore (http://www.christianbenner.com).

#### *Variant annotations*

We analyzed three variant annotations: protein coding (exons), protein non-coding (introns, 5′UTR and 3′UTR), and nonsynonymous SNPs. The latter included transcript ablation, frameshift, stop gained, stop lost, start lost, transcript amplification, inframe insertion, inframe deletion, missense, and protein altering variants. Variants were annotated using the Ensembl Variant Effect Predictor (VEP) ) [7].

#### *Methods of gene-based analysis*

Three regression-based methods based on the summary statistics were applied: SKAT-O [18], PCA [19] and ACAT-V [20]. These methods were implemented in the sumFREGAT R-package (https://cran.r-project.org/web/packages/sumFREGAT/index.html) [21]. The results of different methods were combined using the aggregated Cauchy omnibus test, ACAT-O [20]. The analysis was limited to protein-coding genes with at least two variations that had the estimated summary statistics.

The Bonferroni adjusted significance level for the total number of genes (20,000) was defined as 2.5e-06.

#### *Conditional analysis*

We performed a conditional analysis using the GCTA-COJO tool [22]. The COJO selection procedure was applied to SNPs located within 5 Mb of the gene borders. Conditional summary statistics (p-values, effect estimates) calculated for SNPs within the analyzed gene region were then used as input for gene-based analysis.

### Analysis using exome sequencing data

We performed gene-based association analysis using exome-wide association study (EWAS) data for four chronic pain traits (pain at hip, knee, back and neck) and SGIT of these four traits. Since these EWAS data had smaller sample sizes and lower statistical power in comparison to GWAS data from the main analysis (imputed genotype data from microarrays), we focused only on four musculoskeletal pain types to provide sufficient statistical power for SGIT in SHAHER analysis. We also did not calculate statistics for UGIT due to the lack of statistical power in SHAHER analysis of EWAS data.

#### *Phenotype and exome data used*

To obtain EWAS summary statistics for four pain types we used phenotypic and exome sequencing data from 200,000 Europeans from UK Biobank (a part of the entire European sample used in this study). Chronic pain phenotypes were defined as described above. For exome sequencing data we conducted a quality control according to the guide from https://biobank.ndph.ox.ac.uk/showcase/refer.cgi?id=914. We applied the BCFtools (https://samtools.github.io/bcftools/) to filter SNPs on the multi-sample VCF (data field 23156). The 11,375,237 autosomal SNPs with MAF $< 0.01$ remained after quality control.

#### *Exome-wide association study of pain traits*

The EWAS of each of the four pain traits was carried out using fastGWA-GLMM tool, version 1.94.0 beta [23], --fastGWA-mlm-binary option with filters keeping SNPs with MAF $\geq$ 5e-05 and variant messiness rate $\leq 0.02$. Sex, age, genotyping batch, and the first ten genetic principal components provided by UK Biobank were included as covariates in the analysis to account for fixed effects. We also exploited a sparse genomic relationship matrix in order to correct for random effects attributed to individuals' relationship. We computed this matrix in the entire European sample utilizing fastGWA-GLMM (--make-bK-sparse option, default parameters) for (N = 487,000 with genotyped/imputed common variants). The EWAS results were obtained for GRCh38/ hg38 genomic assembly.

#### *SHAHER analysis*

The SHAHER analysis of EWAS data from four pain types was performed as it described in the 'Materials and Methods' section. We used the phenotypic correlation and genetic correlation matrices from our previous work [1] to estimate linear coefficients for SGIT and obtain EWAS statistics for it. Linear combination coefficients for chronic back, neck, knee and hip pain were 0.46, 0.43, 0.35 and 0.29, respectively.

#### *Gene-based association analysis*

To conduct the gene-based association analysis we calculated a matrix of correlations between every pair of variants within genes using 129,807 unrelated Europeans from UK Biobank utilizing PLINK v2.00a3.7LM (https://www.cog-genomics.org/plink/2.0/) with options --mac 3 --geno 0.02.

We analyzed four variant annotations: LoF, LoF+missense, LoF+protein coding and all intragenic SNPs. The LoF variants include those annotated as frameshift variant, splice acceptor variant, splice donor variant, start loss, stop gain, stop loss, and transcript ablation. The LoF+missense variants additionally include those annotated as transcript amplification, inframe insertion, inframe deletion, missense variant, and protein altering variant. The LoF+protein coding variants additionally include those annotated as synonymous variant, start retained variant, stop retained variant, coding sequence variant, and incomplete terminal codon variant. The all intragenic variants annotation includes all variants within a gene from 5'UTR to 3'UTR.

We used SNPs matching the following criteria: MAF $\leq$ 0.01 and minor allele count (MAC) > 10. The genotypes of ultra-rare variants with MAC $\leq$ 10 were collapsed to a single variant as described in Zhou et al. [24]. Then we calculated the summary statistics for these collapsed variants and used them together with the summary statistics of rare variants. We conducted EWAS for four variant annotations after collapsing. These data were utilized in gene-based analysis with SKAT-O, PCA and ACAT-O tools. All program settings are described above.

#### Polygenic risk score calculation

We calculated the PRSs for SGIT and UGIT using data from entire European sample (discovery and replication samples, $N_{total}$ = 439,762) using three steps algorithm (see Figure SM1).

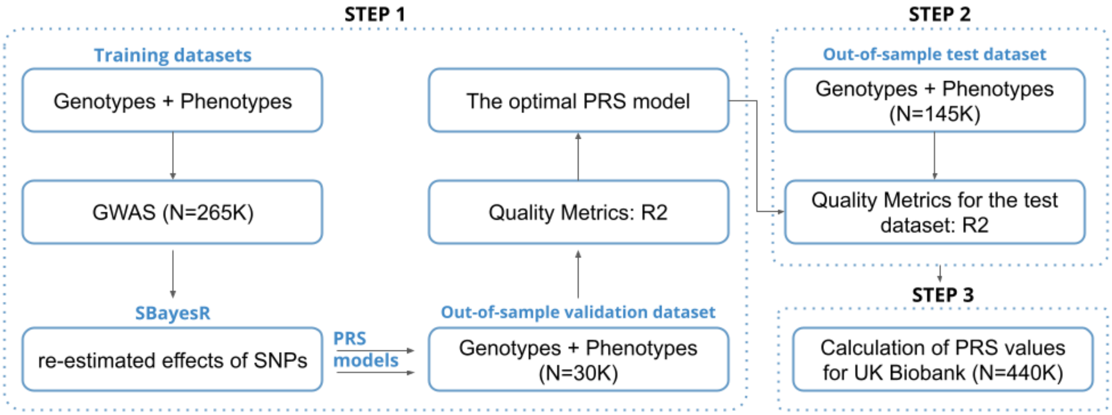


**Figure SM1**. Polygenic risk score calculation scheme.

First, we developed a set of PRS models for each trait using the GWAS for the training set (discovery sample, N = 265,000) and the SBayesR method [25]. SBayesR reweights the effect of each variant according to the marginal estimate of its effect size, statistical strength of association, the degree of correlation between the variant and other variants nearby, and tuning parameters. This method requires a compatible LD matrix file computed using individual-level data from a reference population. For these analyses, we used publicly available shrunk sparse GCTB LD matrix including 1.1 million HapMap3 variants and computed from a random set of 50,000 individuals of European ancestry from the UK Biobank data set. The models were validated in an independent validation dataset of individual-level data (subsample from the European replication sample, N = 30,000), not overlapping with the training sample used for GWAS. In the case of SGIT and UGIT, the prediction accuracy was defined as the proportion of the variance of a phenotype that is explained by PRS values ($R^2$). At the second step, we assessed quality metrics in a test dataset (the other 144,831 Northern Europeans from the European replication sample), which were $R^2$ = 1.8% (95% CI: 1.67-1.95) and $R^2$ = 0.09% (95% CI: 0.06-0.12) for SGIT and UGIT, respectively. At the third step, using the optimal PRS model, we calculated PRS values for every trait in the entire European sample from UK Biobank ($N_{total}$ = 439,762).

### References

1. Tsepilov YA, Freidin MB, Shadrina AS, Sharapov SZ, Elgaeva EE, Zundert J van, et al.

Analysis of genetically independent phenotypes identifies shared genetic factors associated with chronic musculoskeletal pain conditions. Commun Biol. 2020;3. doi:10.1038/s42003-020-1051-9

2. Bulik-Sullivan BK, Loh P-R, Finucane HK, Ripke S, Yang J, Consortium SWG of the PG, et al. LD Score regression distinguishes confounding from polygenicity in genome-wide association studies. Nat Genet. 2015;advance on: 291–295. doi:10.1038/ng.3211

3. Stephens M. A Unified Framework for Association Analysis with Multiple Related Phenotypes. PLoS One. 2013. doi:10.1371/journal.pone.0065245

4. Wei T, Simko V. R package “corrplot”: Visualization of a Correlation Matrix. (Version 0.92). 2021. Available: https://github.com/taiyun/corrplot

5. Boughton AP, Welch RP, Flickinger M, Vandehaar P, Taliun D, Abecasis GR, et al. LocusZoom.js: interactive and embeddable visualization of genetic association study results. doi:10.1093/bioinformatics/btab186

6. Chang CC, Chow CC, Tellier LCAM, Vattikuti S, Purcell SM, Lee JJ. Second-generation PLINK: Rising to the challenge of larger and richer datasets. Gigascience. 2015. doi:10.1186/s13742-015-0047-8

7. McLaren W, Gil L, Hunt SE, Riat HS, Ritchie GRS, Thormann A, et al. The Ensembl Variant Effect Predictor. Genome Biol. 2016. doi:10.1186/s13059-016-0974-4

8. Rogers MF, Shihab HA, Mort M, Cooper DN, Gaunt TR, Campbell C. FATHMM-XF: Accurate prediction of pathogenic point mutations via extended features. Bioinformatics. 2018. doi:10.1093/bioinformatics/btx536

9. Ferlaino M, Rogers MF, Shihab HA, Mort M, Cooper DN, Gaunt TR, et al. An integrative

approach to predicting the functional effects of small indels in non-coding regions of the human genome. BMC Bioinformatics. 2017. doi:10.1186/s12859-017-1862-y

17. Momozawa Y, Dmitrieva J, Théâtre E, Deffontaine V, Rahmouni S, Charloteaux B, et al.

IBD risk loci are enriched in multigenic regulatory modules encompassing putative causative genes. Nat Commun. 2018;9: 1–18. doi:10.1038/s41467-018-04365-8

23. Jiang L, Zheng Z, Fang H, Yang J. Results A generalized linear mixed model association tool for biobank-scale data. doi:10.1038/s41588-021-00954-4

24. Zhou W, Bi W, Zhao Z, Dey KK, Jagadeesh KA, Karczewski KJ, et al. SAIGE-GENE+ improves the efficiency and accuracy of set-based rare variant association tests. Nat Genet. 2022;54. doi:10.1038/s41588-022-01178-w

25. Lloyd-Jones LR, Zeng J, Sidorenko J, Yengo L, Moser G, Kemper KE, et al. Improved polygenic prediction by Bayesian multiple regression on summary statistics.

doi:10.1038/s41467-019-12653-0
