## Supplementary Results for "Decomposing the genetic background of chronic back pain"

### Prioritization of genes in the new associated locus

#### *Literature-based gene prioritization*

Regional association plot for the new rs11079993 locus associated with SGIT is presented on Figure SR1. Three genes were depicted: *CA10* (carbonic anhydrase 10 protein-coding gene), *snoZ178* (small nucleolar RNA Z178) and *LINC01982* (long intergenic non-protein coding RNA 1982).

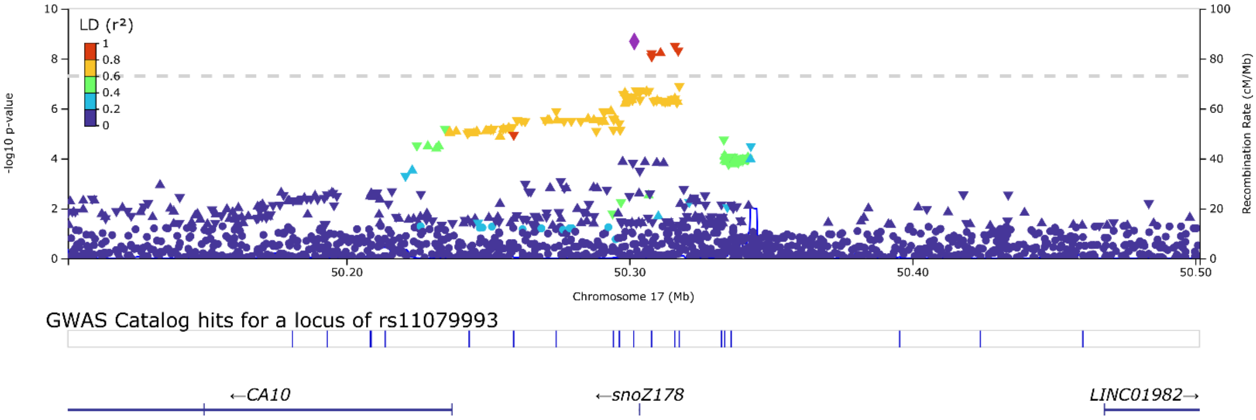


**Figure SR1**. Regional association plot for rs11079993 locus associated with SGIT. The Manhattan plot was created according to the GRCh37 genomic build using the LocusZoom version 0.14 online tool (https://my.locuszoom.org/) [1]. The lead SNP rs11079993 is depicted in purple, blue lines below the Manhattan plot show the location of the nearest genes. Linkage disequilibrium (LD, $r^2$) was calculated automatically using data on Europeans from the 1000 Genomes project with regard to rs12602632 variant ($D'$ = 1.0, $r^2$ = 0.96 with rs11079993 in European population). This variant was used for calculations since the lead SNP was not represented in 1000 Genomes data.

The *CA10* gene encodes a member of the carbonic anhydrase family of zinc metalloenzymes, catalyzing the reversible carbon dioxide hydration. Although the encoded protein does not have a catalytic activity, it is involved in diverse biological processes. For instance, *CA10* is highly expressed in brain and thought to play a role in the central nervous system and brain development [2]. In mice it was shown to be involved in synapse connection [3]. Studies in humans revealed its associations with non-suppurative otitis media and chondroblastoma, which is a rare type of bone cancer. Generally, *CA10* is a tumor suppressor gene, whose promoter methylation is a risk factor for renal cell carcinoma [4], breast cancer [5], and glioma [6]. This gene is also associated with osteoporosis, osteoarthritis and bone mineral density [7,8], airway obstruction, bronchial hyper-reactivity, and inflammation [9]. SNP rs34796998 of the locus was linked to *CA10* in gastroesophageal reflux GWAS [10], and an effect on major depressive disorder was reported for rs34796998 in the same study.

The closest gene to the top SNP of the locus is *snoZ178*. The role of this RNA-coding gene is poorly investigated, but it was recently found in multisite chronic pain GWASs as the nearest gene to the top-associated variant rs11079993 [11,12], though the association had not been replicated to date. The evidence for post-transcriptional RNA modification activity of the encoded nucleolar RNA was shown in other species such as *Schistosoma mansoni* [13].

Most remote from the lead SNP is gene *LINC01982*, which potentially influences carcinogenesis and osteosarcoma tumor development through the immune microenvironment [14]. Furthermore, this gene was revealed in peptic ulcer disease and gastroesophageal reflux cross-trait meta-analysis [15] and shown to be differentially expressed in pheochromocytoma and paraganglioma tumors [16]. The *LINC01982* gene was prioritized in various GWASs on educational attainment, intelligence and cognitive function, neck and shoulder pain, risk-taking behavior, smoking, myocardial infarction, sleep duration, cervical cancer etc. (see https://www.genecards.org/cgi-bin/carddisp.pl?gene=LINC01982&keywords=LINC01982#function for more details).

#### *Prediction of pathogenicity of SNP effects*

The results of variant effect prediction analysis for the rs11079993 locus are presented in Supplementary Tables ST4a, b. In this locus we annotated six variants with three of them attributed to the *snoZ178* small nucleolar RNA gene (rs11079993 was a downstream variant, rs34796998 and rs12602632 were upstream polymorphisms) and the rest of them being intergenic (rs953988, rs12453010 and rs967823). According to the VEP [17] annotation all six SNPs were classified as modifiers, meaning that their effects are hard to predict. FATHMM-XF [18] showed that all of these variants are benign. No insertions or deletions were identified. In addition, according to the VEP build-in literature-based annotation, the rs967823 polymorphism provided association with gastroesophageal reflux and smoking [10] and rs12453010 was linked to neck/shoulder pain in UK Biobank [19].

#### *DEPICT and FUMA gene prioritization*

In DEPICT and FUMA analysis the rs11079993 locus was linked only with the *CA10* gene, although in DEPICT analysis this prioritization was not statistically significant (false discovery rate, FDR $< 0.05$; Supplementary Tables ST5a, b). The FUMA probability of being loss-of-function intolerant [20] (pLI) for this gene was 0.97 meaning that the gene is highly intolerant to loss-of-function mutations (Supplementary Tables ST6a, b). The non-coding residual variation intolerance score [21] (ncRVIS) of *CA10* was estimated as -1.58, indicating its robustness to non-coding variation. Interestingly, the rs967823 variant of the locus was shown to be associated with smoking [22] and educational attainment [23] (Supplementary Table ST6c), which are known risk factors of chronic back pain [24].

#### *Pleiotropic effects on gene expression*

We detected no statistically significant pleiotropic effects of rs11079993 locus on gene expression. The lowest p-value 0.004 of the SMR test was observed for the expression of the *RP11-1018N14.2* gene in the brain spinal cord cervical c-3 tissue.

### References


1. Boughton AP, Welch RP, Flickinger M, Vandehaar P, Taliun D, Abecasis GR, et al. LocusZoom.js: interactive and embeddable visualization of genetic association study

results. doi:10.1093/bioinformatics/btab186

9. Perin P, Potočnik U. Polymorphisms in recent GWA identified asthma genes CA10,

SGK493, and CTNNA3 are associated with disease severity and treatment response in childhood asthma. Immunogenetics. 2014;66. doi:10.1007/s00251-013-0755-0

16. Wang Z, Li Y, Zhong Y, Wang Y, Peng M. Comprehensive analysis of aberrantly expressed competitive endogenous rna network and identification of prognostic biomarkers in pheochromocytoma and paraganglioma. Onco Targets Ther. 2020;13.

doi:10.2147/OTT.S271417
