## Supplementary Figures for "Decomposing the genetic background of chronic back pain"

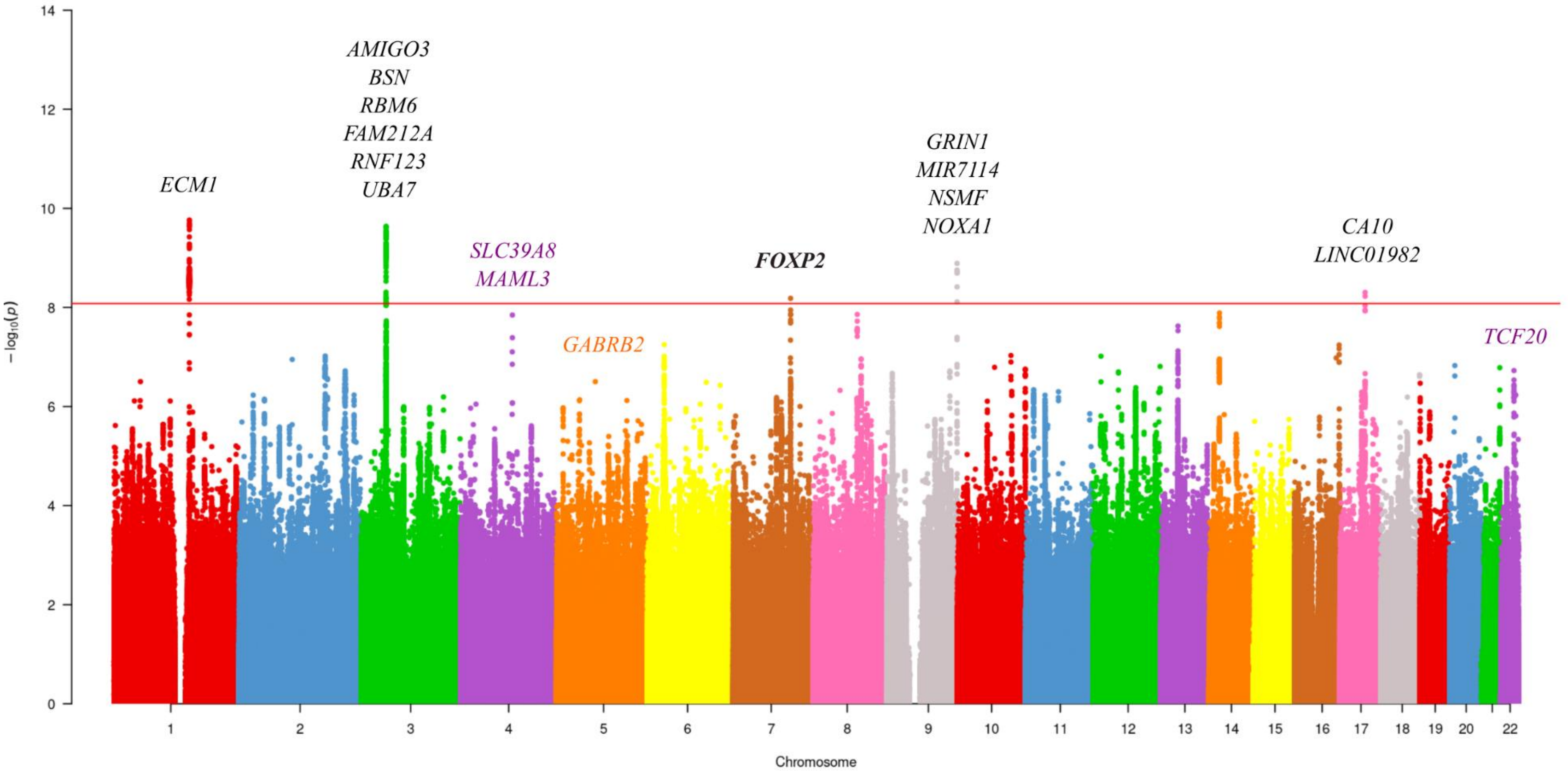


**Supplementary Figure 1**. Manhattan plot for SGIT in discovery sample. The significance threshold was set at p-value < 8.3e-08 (depicted as a horizontal red line). The Manhattan plot was built according to the GRCh37/hg19 genome assembly using variants with minor allele frequency [MAF] ≥ 2e-04. Test statistics were corrected for residual inflation using the LD Score regression intercept 1.04. Genes identified by gene prioritization in the associated loci are written in black. Genes found in gene-based association analysis are written in color of the corresponding chromosome. Gene identified by both approaches is written in bold black.

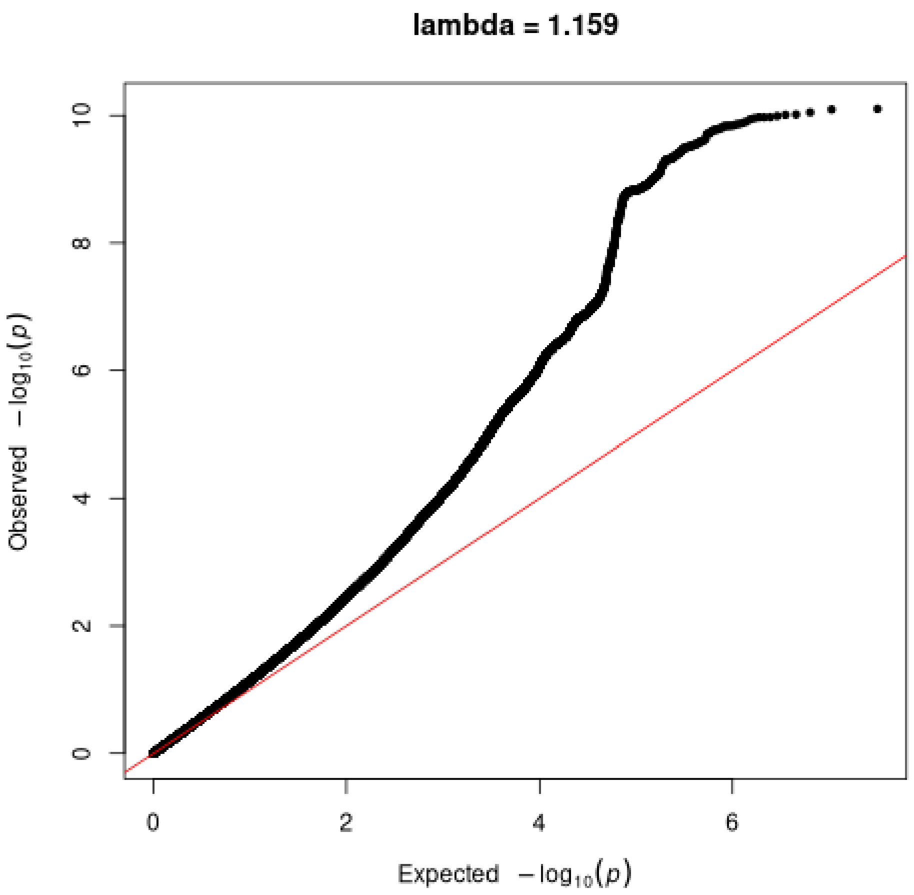


**Supplementary Figure 2**. Quantile-quantile (QQ) plot for SGIT in discovery sample. Lambda of genomic control before correction of the test statistics was 1.16. The plot represents GWAS results without correction for genomic control. No additional filters applied.

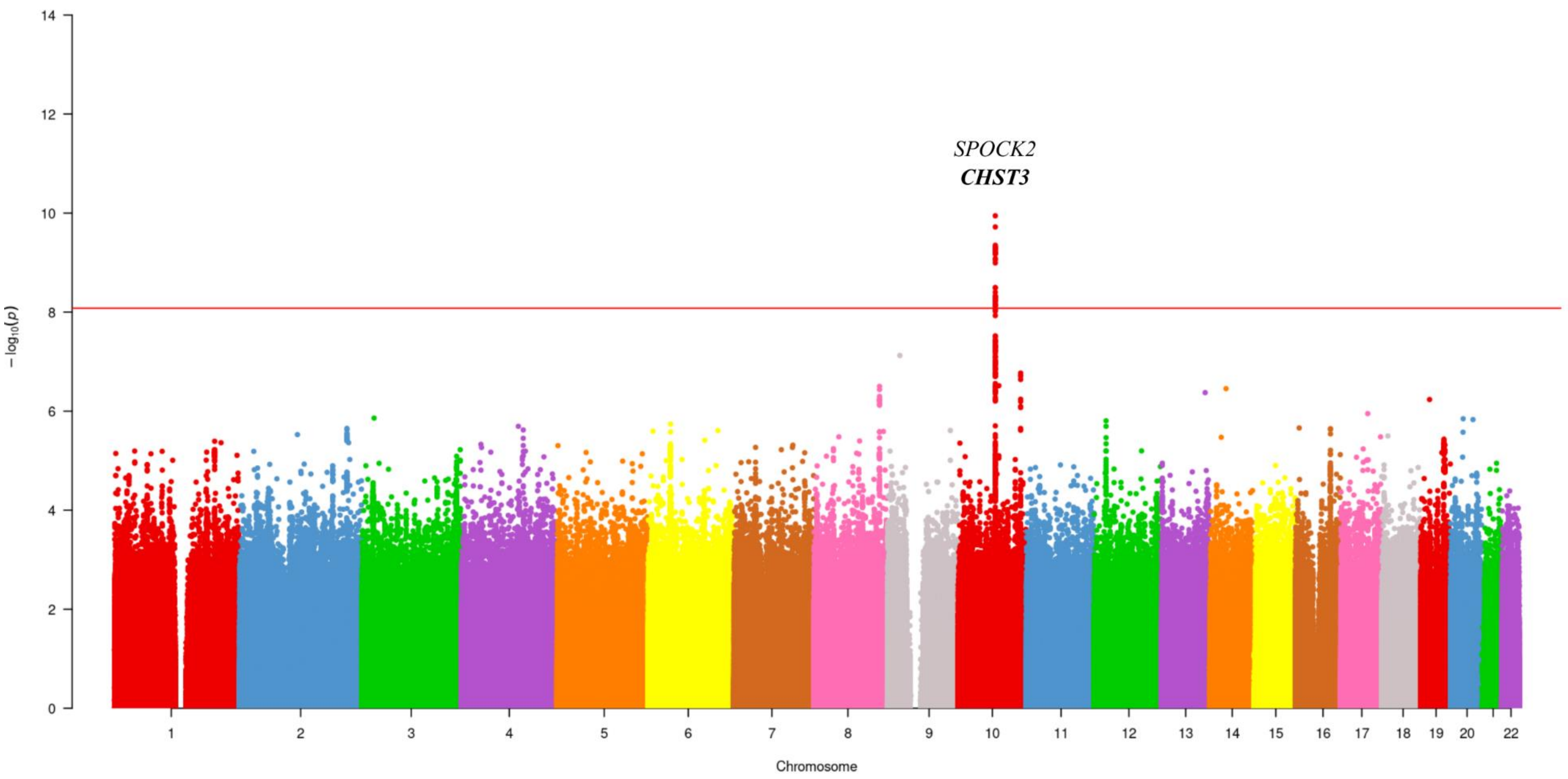


**Supplementary Figure 3**. Manhattan plot for UGIT in discovery sample. The significance threshold was set at p-value < 8.3e-08 (depicted as a horizontal red line). The Manhattan plot was built according to the GRCh37/hg19 genome assembly using variants with minor allele frequency [MAF] ≥ 2e-04. Test statistics were corrected for residual inflation using the LD Score regression intercept 0.99. Genes identified by gene prioritization in the associated loci are written in black. Gene identified by both gene prioritization in the loci and gene-based association analysis is written in bold black.

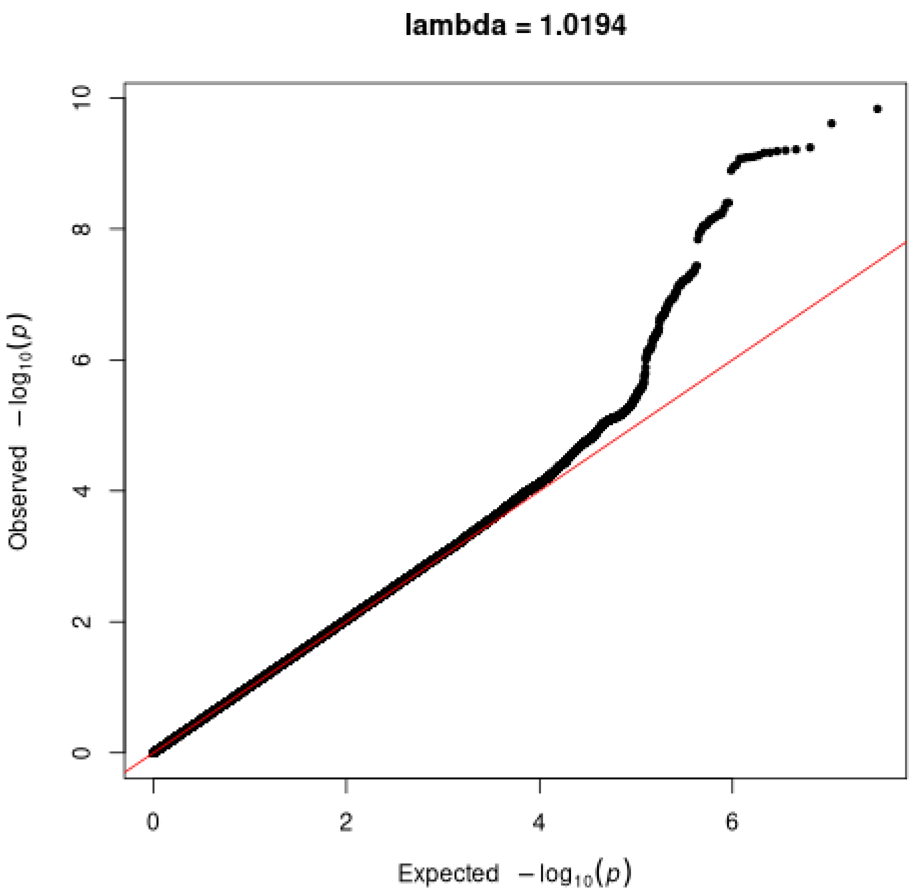


**Supplementary Figure 4**. Quantile-quantile (QQ) plot for UGIT in discovery sample. Lambda of genomic control before correction of the test statistics was 1.02. The plot represents GWAS results without correction for genomic control. No additional filters applied.

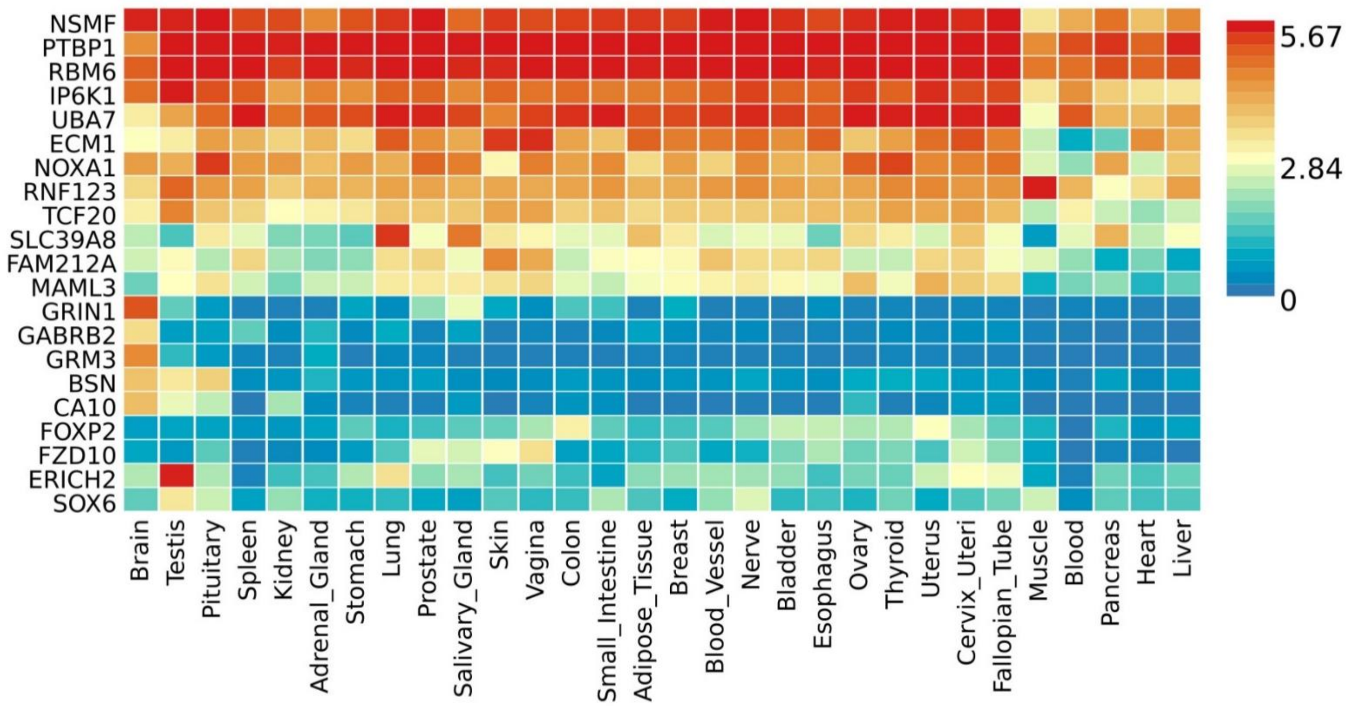


**Supplementary Figure 5**. Gene expression patterns of 23 genes identified for SGIT. Heatmap visualization was prepared using FUMA GENE2FUNC tool and data on GTEx v8 30 general tissue types. As the input data we used: 1) all genes prioritized for SGIT in the GWAS loci (both from the new replicated locus and from the loci previously replicated for GIP1); 2) all genes found in gene-based analysis and replicated in European meta-analysis. Heatmap displays normalized expression value (zero mean normalization of log2 transformed expression), and darker red means higher relative expression of that gene in each label, compared to a darker blue color in the same label. Genes and tissues are ordered by clusters.

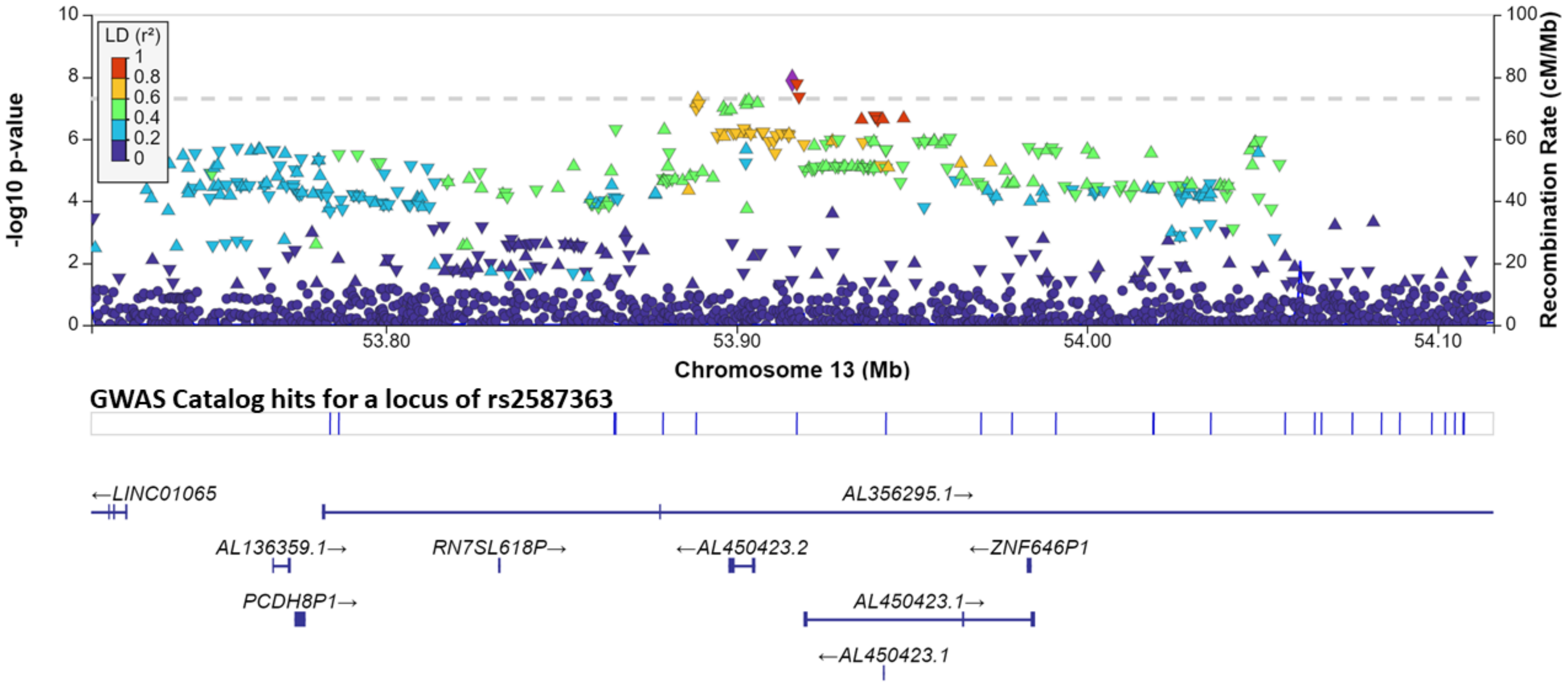


**Supplementary Figure 6**. Regional association plot for rs2587363 locus associated with SGIT under suggestive significance threshold p-value < 8.3e-08. The Manhattan plot was created according to the GRCh37/hg19 genomic build using LocusZoom version 0.14 online tool (https://my.locuszoom.org/). Only genetic variants with minor allele frequency [MAF] ≥ 2e-04 are presented. Test statistics were corrected for residual inflation using the LD Score regression intercept 1.04. The lead SNP rs2587363 is depicted in purple, blue lines below the Manhattan plot show the location of the nearest genes. Linkage disequilibrium (LD, $r^2$) was calculated automatically using data on Europeans from 1000 Genomes project with regard to the lead SNP.
