## Supplementary Tables for "Decomposing the genetic background of chronic back pain": Supplementary Table ST1. Description of the sample.docx

**Supplementary Table ST1**. Description of the study sample

|  | Prevalence in a sample | Sample size | Age (mean ± SD), *years* | BMI (mean ± SD), *kg/m^2^* | Women, *%* |
| --- | --- | --- | --- | --- | --- |
| **Discovery sample (N = 265,000)** | | | | | |
| Chronic back pain | 17.9% | Cases (N = 47,507) | 57.65 (7.99) | 28.33 (5.18) | 45.33 |
|  |  | Controls (N = 217,493) | 57.26 (8.03) | 27.15 (4.61) | 45.96 |
| Chronic neck pain | 16.3% | Cases (N = 43,287) | 57.73 (7.79) | 27.90 (5.02) | 40.98 |
|  |  | Controls (N = 221,713) | 57.25 (8.07) | 27.25 (4.68) | 46.80 |
| Chronic hip pain | 9.2% | Cases (N = 24,300) | 59.15 (7.44) | 28.91 (5.40) | 36.59 |
|  |  | Controls (N = 240,700) | 57.15 (8.06) | 27.20 (4.64) | 46.79 |
| Chronic knee pain | 17.5% | Cases (N = 46,292) | 58.61 (7.59) | 29.18 (5.37) | 46.83 |
|  |  | Controls (N = 218,708) | 57.06 (8.09) | 26.97 (4.50) | 45.64 |
| Chronic stomach pain | 4.8% | Cases (N = 12,739) | 55.90 (8.23) | 27.93 (5.39) | 37.53 |
|  |  | Controls (N = 252,261) | 57.40 (8.01) | 27.33 (4.70) | 46.27 |
| Chronic headache | 9.2% | Cases (N = 24,421) | 55.00 (7.85) | 27.33 (5.11) | 29.56 |
|  |  | Controls (N = 240,579) | 57.57 (8.01) | 27.36 (4.70) | 47.50 |
| **Replication sample (N = 191,580)** | | | | | |
| ***African ancestry sample* (N = 7,541)** | | | | | |
| Chronic back pain | 21.0% | Cases (N = 1,586) | 53.77 (8.24) | 30.62 (5.79) | 37.01 |
|  |  | Controls (N = 5,955) | 52.04 (8.00) | 29.27 (5.13) | 44.25 |
| Chronic neck pain | 16.1% | Cases (N = 1,217) | 54.38 (7.98) | 30.06 (5.52) | 34.92 |
|  |  | Controls (N = 6,324) | 52.02 (8.04) | 29.45 (5.25) | 44.23 |
| Chronic hip pain | 8.5% | Cases (N = 641) | 55.00 (7.91) | 31.30 (6.14) | 31.05 |
|  |  | Controls (N = 6,900) | 52.16 (8.05) | 29.39 (5.19) | 43.81 |
| Chronic knee pain | 20.4% | Cases (N = 1,539) | 54.67 (8.30) | 31.64 (6.11) | 31.71 |
|  |  | Controls (N = 6,002) | 51.82 (7.92) | 29.01 (4.93) | 45.55 |
| Chronic stomach pain | 6.7% | Cases (N = 508) | 50.90 (7.64) | 29.92 (5.70) | 31.89 |
|  |  | Controls (N = 7,033) | 52.51 (8.10) | 29.52 (5.27) | 43.51 |
| Chronic headache | 8.1% | Cases (N = 610) | 50.56 (7.37) | 30.27 (5.65) | 30.33 |
|  |  | Controls (N = 6,931) | 52.56 (8.12) | 29.49 (5.26) | 43.82 |
| ***European ancestry sample* (N = 174,831)** | | | | | |
| Chronic back pain | 18.0% | Cases (N = 31,428) | 57.62 (7.96) | 28.36 (5.22) | 45.09 |
|  |  | Controls (N = 143,403) | 57.26 (8.02) | 27.14 (4.58) | 46.31 |
| Chronic neck pain | 16.3% | Cases (N = 28,482) | 57.82 (7.76) | 27.92 (5.02) | 40.92 |
|  |  | Controls (N = 146,349) | 57.23 (8.06) | 27.25 (4.66) | 47.09 |
| Chronic hip pain | 9.2% | Cases (N = 16,022) | 59.26 (7.40) | 28.86 (5.41) | 36.66 |
|  |  | Controls (N = 158,809) | 57.13 (8.05) | 27.21 (4.63) | 47.04 |
| Chronic knee pain | 17.3% | Cases (N = 30,173) | 58.71 (7.54) | 29.24 (5.41) | 47.01 |
|  |  | Controls (N = 144,658) | 57.04 (8.08) | 26.97 (4.47) | 45.89 |
| Chronic stomach pain | 4.9% | Cases (N = 8,465) | 56.05 (8.20) | 27.96 (5.43) | 38.35 |
|  |  | Controls (N = 166,366) | 57.39 (8.00) | 27.33 (4.69) | 46.48 |
| Chronic headache | 9.2% | Cases (N = 16,101) | 55.14 (7.73) | 27.35 (5.12) | 28.98 |
|  |  | Controls (N = 158,730) | 57.55 (8.01) | 27.36 (4.68) | 47.82 |
| ***South Asian ancestry sample* (N = 9,208)** | | | | | |
| Chronic back pain | 21.6% | Cases (N = 1,993) | 54.66 (8.51) | 27.76 (4.58) | 49.62 |
|  |  | Controls (N = 7,215) | 53.87 (8.47) | 26.92 (4.23) | 54.66 |
| Chronic neck pain | 20.2% | Cases (N = 1,864) | 54.65 (8.24) | 27.43 (4.56) | 43.13 |
|  |  | Controls (N = 7,344) | 53.88 (8.53) | 27.01 (4.25) | 56.22 |
| Chronic hip pain | 6.6% | Cases (N = 610) | 56.61 (8.21) | 28.30 (4.90) | 44.26 |
|  |  | Controls (N = 8,598) | 53.86 (8.47) | 27.01 (4.26) | 54.23 |
| Chronic knee pain | 20.1% | Cases (N = 1,850) | 55.97 (8.23) | 28.52 (4.86) | 44.54 |
|  |  | Controls (N = 7,358) | 53.55 (8.47) | 26.74 (4.09) | 55.84 |
| Chronic stomach pain | 5.3% | Cases (N = 488) | 52.83 (7.98) | 27.64 (5.33) | 43.44 |
|  |  | Controls (N = 8,720) | 54.11 (8.50) | 27.07 (4.25) | 54.14 |
| Chronic headache | 10.6% | Cases (N = 975) | 51.45 (7.59) | 27.65 (4.94) | 35.18 |
|  |  | Controls (N = 8,233) | 54.34 (8.53) | 27.03 (4.24) | 55.75 |
| Chronic hip pain | 8.5% | Cases (N = 641) | 55.00 (7.91) | 31.30 (6.14) | 31.05 |
|  |  | Controls (N = 6,900) | 52.16 (8.05) | 29.39 (5.19) | 43.81 |

BMI – body mass index; N – sample size; SD – standard deviation
